# Pathways through which water, sanitation, hygiene, and nutrition interventions reduce antibiotic use in young children: a mediation analysis of a cluster-randomized trial

**DOI:** 10.1101/2024.10.13.24315425

**Authors:** Anna Nguyen, Gabby Barratt Heitmann, Andrew Mertens, Sania Ashraf, Md Ziaur Rahman, Shahjahan Ali, Mahbub Rahman, Benjamin F. Arnold, Jessica A. Grembi, Audrie Lin, Ayse Ercumen, Jade Benjamin-Chung

**Affiliations:** Department of Epidemiology and Population Health, School of Medicine, Stanford University, Stanford, CA, USA; Division of Epidemiology and Biostatistics, School of Public Health, University of California, Berkeley, Berkeley, CA, USA; Environmental Interventions Unit, Health System and Population Studies Division, icddr,b, Dhaka 1212, Bangladesh; Department of Microbiology and Environmental Toxicology, UC Santa Cruz, Santa Cruz, CA, USA; Infectious Disease Division, International Centre for Diarrhoeal Disease Research, Bangladesh, Dhaka, Bangladesh; Francis I. Proctor Foundation and Department of Ophthalmology, University of California, San Francisco, San Francisco, CA, USA; Division of Infectious Diseases and Geographic Medicine, Department of Medicine, School of Medicine, Stanford University, Stanford, CA, USA; Department of Forestry and Environmental Resources, North Carolina State University, Raleigh, NC, USA; Chan Zuckerberg Biohub, San Francisco, CA, USA

**Author notes:** Corresponding author: Jade Benjamin-Chung.

## Abstract

**Background:** Low-cost, household-level water, sanitation, and hygiene (WASH) and nutrition interventions can reduce pediatric antibiotic use, but the mechanism through which interventions reduce antibiotic use has not been investigated.

**Methods:** We conducted a causal mediation analysis using data from the WASH Benefits Bangladesh cluster-randomized trial (NCT01590095). Among a subsample of children within the WSH, nutrition, nutrition+WSH, and controls arms (N=1,409), we recorded caregiver-reported antibiotic use at ages 14 and 28 months and collected stool at age 14 months. Mediators included caregiver-reported child diarrhea, acute respiratory infection (ARI), and fever; and enteric pathogen carriage in stool measured by qPCR. Models controlled for mediator-outcome confounders.

**Findings:** The receipt of any WSH or nutrition intervention reduced antibiotic use in the past month by 5.5 percentage points (95% CI 1.2, 9.9) through all pathways, from 49.5% (95% CI 45.9%, 53.0%) in the control group to 45.0 % (95% CI 42.7%, 47.2%) in the pooled intervention group. Interventions reduced antibiotic use by 0.6 percentage points (95% CI 0.1, 1.3) through reduced diarrhea, 0.7 percentage points (95% CI 0.1, 1.5) through reduced ARI with fever, and 1.8 percentage points (95% CI 0.5, 3.5) through reduced prevalence of enteric viruses. Interventions reduced antibiotic use through any mediator by 2.5 percentage points (95% CI 0.2, 5.3).

**Interpretation:** Our findings bolster a causal interpretation that WASH and nutrition interventions reduced pediatric antibiotic use through reduced infections in a rural, low-income population.

**Funding:** Bill & Melinda Gates Foundation, National Institute of Allergy and Infectious Diseases

**Research in Context:** *Evidence before this study:* We searched for primary studies and systematic reviews that investigated mediation of antibiotic use by water, sanitation and hygiene interventions in Scopus using (TITLE-ABS-KEY((“WASH” OR “sanitation” OR “water” OR “hygiene” OR “nutrition”) AND (“antibiot*”) AND (“interven*”) AND (“mediat*” OR “indirect effect*” OR “pathway” OR “mechanism”) AND (“use” OR “practice*”)). We included all publications until September 4, 2024. We restricted results to studies in English, focused on humans, and within medicine, agricultural and biological sciences, immunology or microbiology, or environmental science. Our search yielded 115 studies. We found no relevant research studies. Four review studies discussed the need for improved sanitation and drinking water as an AMR control strategy in LMICs. Two study protocols described longitudinal observational studies in LMICs that will explore the relationship between WASH and antibiotic resistance.

*Added value of this study:* We used causal mediation analysis to investigate mechanisms through which WASH and nutrition interventions reduced antibiotic use in young children in a community setting in rural Bangladesh. This study is rigorous because it leverages a randomized trial with high intervention adherence and includes objectively measured mediators. We found that WASH and nutrition interventions reduced antibiotic use via reduced diarrhea, ARI with fever, and enteric virus carriage. This study improves on previous studies by identifying a specific mechanism through which WASH and nutrition interventions reduced pediatric antibiotic use in an understudied setting and population.

*Implications of all the available evidence:* In a previous analysis of a randomized trial of WASH and nutrition interventions, we found that pediatric antibiotic use was lower in the intervention arms compared to control. Here, using causal mediation analysis, we identified several biologically plausible pathways through which interventions likely reduced antibiotic use. This analysis bolsters a causal interpretation that low-cost, household-level WASH and nutrition interventions can reduce pediatric antibiotic use in settings with similar infectious disease dynamics and antimicrobial access.

## Introduction

Antimicrobial resistance (AMR) is one of the top global health threats and was linked to 4.95 million deaths worldwide in 2019^1^, with a larger burden than malaria or HIV. Deaths and antimicrobial-resistant infections are concentrated among low- and middle-income countries (LMICs)^1^, where there are high rates of antibiotic misuse due to inappropriate prescriptions for non-bacterial infections^2,3^. Diarrhea cases of unconfirmed etiology in LMICs are frequently treated with antibiotics^4^, despite enteric viruses being a leading cause of diarrhea among children under 24 months.^5,6^ Consumption of antibiotics that have higher potential for resistance according to the WHO grew by 165% in LMICs from 2000-2015^7^.

Though much research has investigated AMR transmission in hospital settings^8^, a growing body of research points to a high burden of AMR in community settings in LMICs^9^. Antibiotic resistant pathogens can be spread through contaminated water, food, or fomites^10^. Improved WASH infrastructure could reduce transmission of antibiotic resistant pathogens from human and zoonotic sources^11^. Improved WASH was found to reduce the abundance of antibiotic resistance genes (ARGs) by 22% in community settings^12^ and a modeling analysis suggested that universal access to improved WASH could reduce the number of antibiotics used by 60% in LMICs^13^. Enhanced child nutrition may also play a role in combating AMR by boosting immunity and prevent enteric and respiratory infections^14^, which are drivers of antibiotic use and AMR^3,15^.

We previously found that low-cost, household-level WASH and nutrition interventions reduced antibiotic use in rural communities in Bangladesh by 10% and 14% within the WASH Benefits cluster-randomized trial^16^. However, the mechanism for these reductions is poorly understood. It is possible that WASH and nutrition interventions reduce diarrhea, enteric pathogen infections, and respiratory illnesses, and these reductions resulted in lower antibiotic use. Among children under 2 years, WASH interventions reduced diarrhea by 31-40%^17^, enteric virus carriage by approximately 50%^18^, and acute respiratory illness (ARI) by 25-33%^19^. Nutrition interventions reduced childhood diarrhea by 26%^17^ and enteric virus carriage by up to 42%^18^. No prior studies have formally investigated pathways through which WASH interventions reduce antibiotic use in LMICs.

We investigated whether reductions in pediatric antibiotic use due to WASH and nutrition interventions were mediated by reduced enteric virus carriage, diarrhea, or ARI in children under 2 years in rural Bangladesh using data from the WASH Benefits Bangladesh trial^17^. Understanding the mechanisms through which WASH and nutrition interventions reduce antibiotic use will inform future health interventions to reduce unnecessary antibiotic use, and in turn, reduce AMR.

## Methods

We published a pre-analysis plan at https://osf.io/ytmcr and listed deviations from this plan in Supplement 1.

### Study design & population

We analyzed data from the WASH Benefits Bangladesh trial, which enrolled 720 clusters including 5,551 pregnant mothers in four districts of rural Bangladesh between May 31, 2012, and July 7, 2013^17^. Clusters were randomly allocated to the following arms: water (W), hygiene (H), sanitation (S), combined water, sanitation, and hygiene (WSH), nutrition (N), and combined N+WSH. Randomization was stratified by geographic block. Index children born to pregnant mothers were followed for two years. The primary outcomes were child diarrhea and growth.

This study utilized data from the environmental enteric dysfunction (EED) substudy,of WASH Benefits in which additional measurements were collected in 267 clusters in the N, WSH, N+WSH, and C arms.^20^ Clusters were equally distributed between study arms but did not adhere to the geographic matching of the parent trial. EED cohort enrolled 1,531 children at age 14 months and 1,531 children at age 28 months. Our analysis was restricted to children with non-missing antibiotic use in the follow-up rounds when they were approximately 14 months or 28 months (N = 1,528) old. Antibiotic use data was available for >99% of children at both 14 and 28 months.

### Interventions

The WSH intervention included chlorine tablets for water treatment and a safe water storage vessel; double-pit latrine upgrades, child potties, and hoes for removing feces; and handwashing stations. The nutrition intervention included promotion of age-appropriate maternal and infant nutrition practices and lipid-based nutrient (LNS) supplements for children from 6-24 months of age. Interventions were given to participants free of charge, and consumables were restocked during trial follow-up. Intervention fidelity was high^21^. Additional intervention details are included in Supplement 2.

### Mediators

Mediators included caregiver-reported diarrhea, acute respiratory infection (ARI), ARI with fever, difficulty breathing in the prior 7 days, fever in the prior 7- or 14-days, and enteric virus carriage. Diarrhea was defined as three or more loose or watery stools within 24 hours or at least one bloody stool in the prior 7 days^22^, ARI was defined as persistent cough or panting/wheezing/difficulty breathing in the prior 7 days^23^. Diarrhea, ARI, difficulty breathing, and fever in the prior 7-days were measured at approximately 14 and 28 months, while fever in the prior 14-days was only measured at approximately 28 months. Stool samples were collected at age 14 months and analyzed for enteropathogens via quantitative polymerase chain reaction (PCR) with a Taqman array card^5,24^. Further details of enteropathogen data collection have been described elsewhere^18^. The Taqman enteropathogen panel included viruses, bacteria, and protozoa, but we only included carriage of enteric viruses as a mediator because a prior analysis found no effects of interventions on bacteria or protozoa^18^. Viruses included in the panel include adenovirus 40/41, norovirus GI, norovirus GII, sapovirus, rotavirus, and astrovirus. Here, we focused on adenovirus 40/41, norovirus GII, and sapovirus carriage as potential mediators, as prior analysis of this data did not find that interventions had an effect on the other viruses.^18^ We also included indicators for combinations of mediators, including any enteric virus that is included in the Taqman panel, any enteric virus with diarrhea, and any mediator (diarrhea, fever, ARI, or enteric virus).

### Outcomes

Our primary outcome was any caregiver-reported antibiotic use by index children within the past 30 or 90 days measured at age 14 and 28 months. Field staff provided caregivers with a list of commonly used antibiotics^16^ and asked them to report the index child’s antibiotic use. These antibiotics included Ceftriaxone, Cefixime, Amoxycillin, Ciprofloxacin, Metronidazole, Cefuroxime, Azithromycin, Cloxacillin, Ampicillin, Flucloxacillin, Levofloxacin, Gentamicin, Cephradine, Cefepime, Erythromycin, Neomycin, and Betapen. For mediation by diarrhea, ARI, and fever, which used a 7- or 14-day recall period, we used antibiotic use within the past 30 days and coded children with the last reported antibiotic use more than 30 days before symptom measurements as not having taken antibiotics. For mediation by enteric virus carriage, we conducted analyses using recall periods of 30 and 90 days for antibiotic use because enteric viruses can persist in stool for up to 12 months^25,26^. We conducted additional analyses on the secondary outcomes of multiple antibiotic use, number of episodes of antibiotic use, and number of days of antibiotic use in the 90-day recall period.

### Statistical analysis

Our primary analysis compared a pooled intervention group (WSH, N, or N+WSH) to the control arm. We conducted secondary analyses comparing individual intervention arms to the control arm, as well as those that grouped (1) WSH and WSH + Nutrition and (2) Nutrition and WSH + Nutrition. We modeled treatment-outcome, treatment-mediator, and mediator-outcome relationships using generalized linear models with robust standard errors that account for clustering at the block level. For continuous outcomes, we used a Gaussian family and identity link function, and for categorical outcomes, we used a Poisson family and log link function. We estimated prevalence differences from treatment-outcome models using g-computation with logistic regression and constructed confidence intervals with 1,000 iterations of bootstrapped resampling at the block level. All models were adjusted for child age to account for multiple measurements taken at 14 and 28 months for each child.

We adjusted mediator-outcome models for any of the potential confounders with a likelihood ratio test p-value < 0.2: time of fecal sample/data collection (measured in 3-month periods); sex, and birth order; mother’s age, height and education; household food insecurity; number of individuals <18 years in household; number of individuals living in compound; distance to household’s primary drinking water source; housing materials; and a household wealth index calculated from the first principal component of a principal components analysis of household assets. All covariates were measured at baseline except for month of measurement, child age, and child sex.

The total effect of WASH and nutrition interventions on antibiotic use can be decomposed into the mediated effect (i.e, “natural indirect effect” (NIE)) or the direct effect (i.e., “natural direct effect”)^27^. The NIE is the difference in potential outcomes under the predicted values of the mediator if all children had been in the treatment arm versus if all children had been in the control arm. We assumed that a NIE was only possible for a particular mediator if there was a significant mediator-outcome relationship, independent from the intervention. For mediators with significant mediator-outcome relationships (p-value < 0.05), we estimated NIEs and 95% confidence intervals using a quasi-Bayesian Monte Carlo approach with 1,000 simulations as implemented by the mediation R package^24^. In these mediation analyses, we used intervention-mediator and mediator-outcome models that followed the same generalized linear modeling approach described above. We adjusted both models for mediator-outcome confounders that were identified in covariate screening. We assessed potential intervention-mediator interactions within mediation models. For any models in which there was a difference in NIE in the treatment vs control greater than 1% or t-test p-value < 0.2 when a treatment-mediator interaction term was included, we reported the NIEs estimated while holding either treatment status constant to treated (the “total” NIE) or control (the “pure” NIE) (additional details in Supplement 3).^28^

Analyses were conducted in R (version 4.1.3). We have published replication materials on Open Science Framework (https://osf.io/ytmcr)

### Assumptions of mediation analyses

Because our analysis uses data from a randomized trial with baseline balance, we reasonably met the assumption that there is no unmeasured confounding of the treatment-outcome relationship. We attempted to meet the assumption of no mediator-outcome confounding through covariate adjustment (Table S1), although residual unmeasured confounding is always a possibility. We performed a complete case analysis, assuming missing data are missing completely at random.

### Sensitivity analysis

Because some children may carry enteric viruses at low levels, we performed a sensitivity analysis of mediation by enteric viruses in children whose pathogen loads reflect diarrheal etiology. We used published Ct cutoff values for adenovirus 40/41, norovirus GII and sapovirus from the MAL-ED study^29^. We also conducted a negative control analysis using caregiver-reported bruising in the prior 7 days to detect potential mediator misclassification or residual unmeasured confounding.^30,31^

#### Ethics statement

All study participants gave written informed consent in Bengali. This study protocol was approved for human subjects research at the icddr,b (PR-11063), University of California, Berkeley (2011-09-3652), and Stanford University (25863).

## Results

This analysis included 1,716 children present at 14 or 28 month follow-ups (N=3,056 measurements) (Figure S1). Baseline characteristics, including child sex, mother’s age, and household materials, were well-balanced at baseline between study arms (Table 1, Table S2). In all arms, over two thirds of participants were food secure, while over half of mothers had obtained a secondary education. At 14 months, 128 (5.9%) children were missing stool samples in the intervention arm, and 74 (10.7%) were missing a stool sample in the control arm. Generally, diarrhea, ARI, and enteric virus carriage were more prevalent at 14 months compared to 28 months (Table 2, Table S3). Co-prevalence of diarrhea, ARI, and enteric virus carriage was relatively rare; <1% of children experienced diarrhea and ARI concurrently and 2% of children experienced ARI while carrying an enteric virus (Figure S2, Table S4).

**Table 1:**
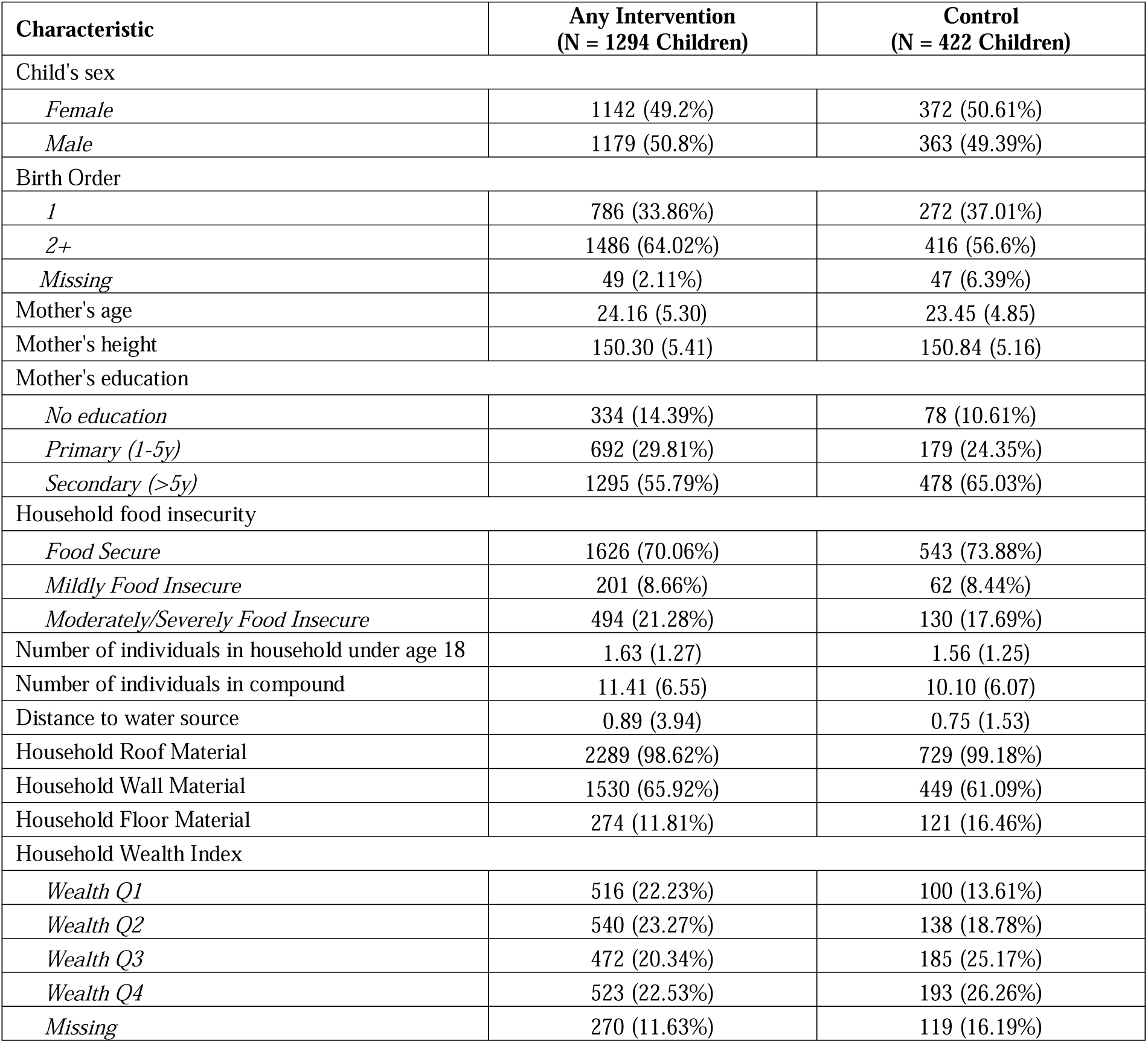
Distribution of Baseline Characteristics by Treatment Group. Sample sizes, child demographics, and household characteristics, in the pooled intervention vs. control arm. For categorical variables, the number of occurrences and percentages are reported. For continuous variables, the mean and standard deviation (SD) are reported. We report the distribution of these variables in individual, unpooled treatment arms in Supplemental Table S2.

| Characteristic | Any Intervention<br>(N = 1294 Children) | Control<br>(N = 422 Children) |
| --- | --- | --- |
| Child's sex |  |  |
| *Female* | 1142 (49.2%) | 372 (50.61%) |
| *Male* | 1179 (50.8%) | 363 (49.39%) |
| Birth Order |  |  |
| *1* | 786 (33.86%) | 272 (37.01%) |
| *2+* | 1486 (64.02%) | 416 (56.6%) |
| *Missing* | 49 (2.11%) | 47 (6.39%) |
| Mother's age | 24.16 (5.30) | 23.45 (4.85) |
| Mother's height | 150.30 (5.41) | 150.84 (5.16) |
| Mother's education |  |  |
| *No education* | 334 (14.39%) | 78 (10.61%) |
| *Primary (1-5y)* | 692 (29.81%) | 179 (24.35%) |
| *Secondary (>5y)* | 1295 (55.79%) | 478 (65.03%) |
| Household food insecurity |  |  |
| *Food Secure* | 1626 (70.06%) | 543 (73.88%) |
| *Mildly Food Insecure* | 201 (8.66%) | 62 (8.44%) |
| *Moderately/Severely Food Insecure* | 494 (21.28%) | 130 (17.69%) |
| Number of individuals in household under age 18 | 1.63 (1.27) | 1.56 (1.25) |
| Number of individuals in compound | 11.41 (6.55) | 10.10 (6.07) |
| Distance to water source | 0.89 (3.94) | 0.75 (1.53) |
| Household Roof Material | 2289 (98.62%) | 729 (99.18%) |
| Household Wall Material | 1530 (65.92%) | 449 (61.09%) |
| Household Floor Material | 274 (11.81%) | 121 (16.46%) |
| Household Wealth Index |  |  |
| *Wealth Q1* | 516 (22.23%) | 100 (13.61%) |
| *Wealth Q2* | 540 (23.27%) | 138 (18.78%) |
| *Wealth Q3* | 472 (20.34%) | 185 (25.17%) |
| *Wealth Q4* | 523 (22.53%) | 193 (26.26%) |
| *Missing* | 270 (11.63%) | 119 (16.19%) |

**Table 2:**
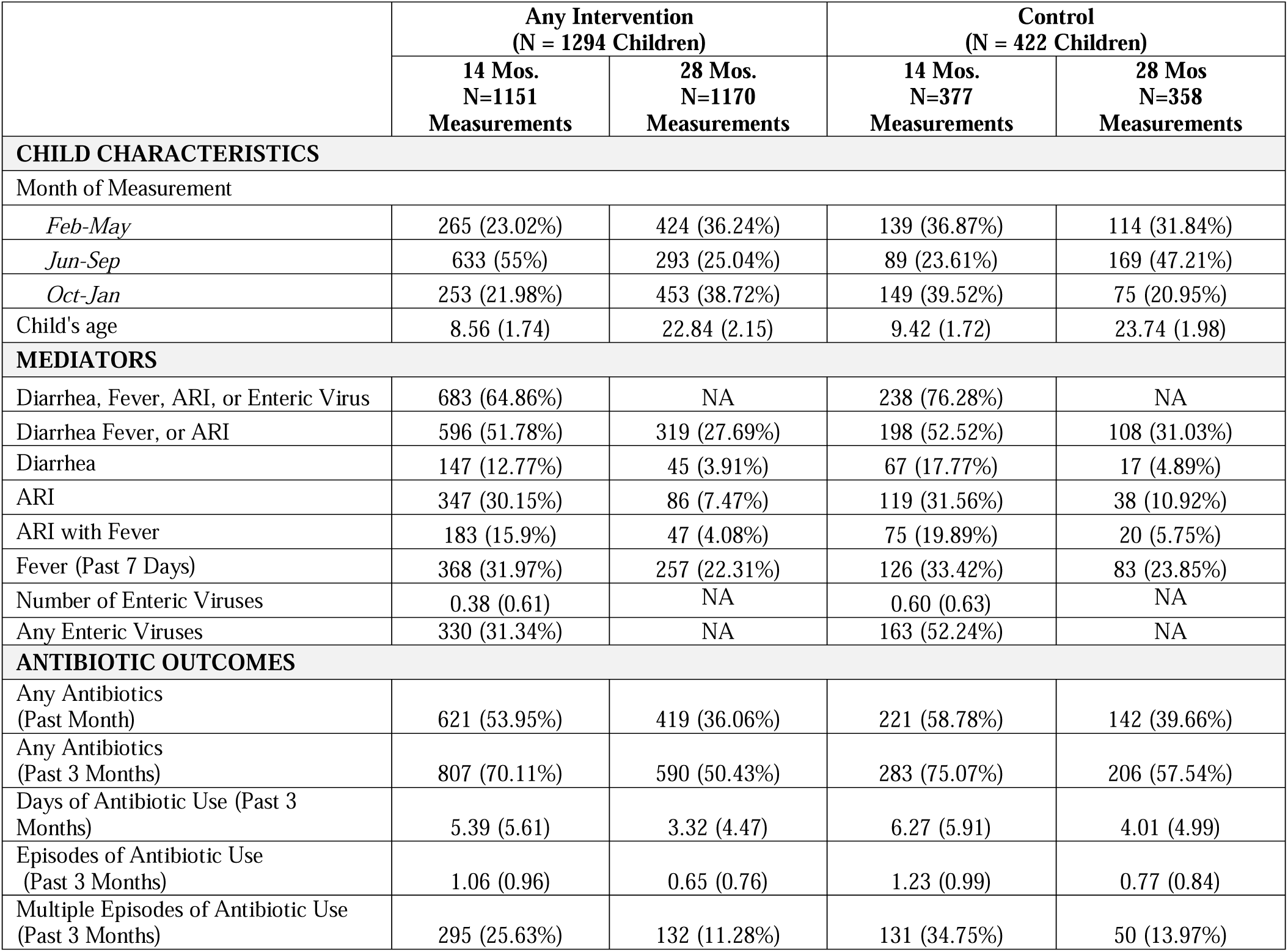
Prevalence of Mediators and Antibiotic Use Outcomes by Treatment Group at 14- and 28-months follow-up. Sample sizes, child characteristics, mediator prevalence, and antibiotic use prevalence, in the pooled intervention vs. control arm by follow-up period. For categorical variables, the number of occurrences and percentages are reported. For continuous variables, the mean and standard deviation (SD) are reported.

**Table 3:**
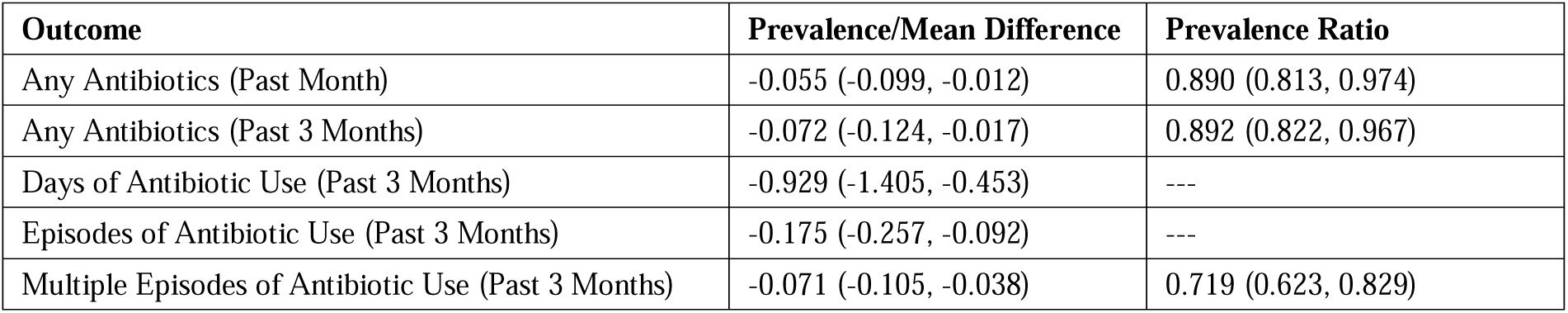
Total Effects of Any WASH or Nutrition Intervention on Antibiotic Use. Point estimates and 95% confidence intervals (CIs) for the total effect of any WASH or nutrition intervention on antibiotic use outcomes. For categorical outcomes (any antibiotic use, multiple episodes of antibiotic use), the prevalence difference and prevalence ratio are reported. For continuous outcomes (days of antibiotic use, episodes of antibiotic use), only the mean difference is reported.

| Outcome | Prevalence/Mean Difference | Prevalence Ratio |
| --- | --- | --- |
| Any Antibiotics (Past Month) | -0.055 (-0.099, -0.012) | 0.890 (0.813, 0.974) |
| Any Antibiotics (Past 3 Months) | -0.072 (-0.124, -0.017) | 0.892 (0.822, 0.967) |
| Days of Antibiotic Use (Past 3 Months) | -0.929 (-1.405, -0.453) | --- |
| Episodes of Antibiotic Use (Past 3 Months) | -0.175 (-0.257, -0.092) | --- |
| Multiple Episodes of Antibiotic Use (Past 3 Months) | -0.071 (-0.105, -0.038) | 0.719 (0.623, 0.829) |

### Total effects

WSH and nutrition interventions (henceforth, “interventions”) led to a modest reduction in antibiotic use, and the strength of evidence was strongest for the effect of interventions on multiple episodes of antibiotic use in the past 3 months (Figure S3, Table S5). Interventions reduced the prevalence of any antibiotic use in the past month from 50% (95% CI 46%, 53%) in the control arm to 45% (95% CI 43%, 47%) in the pooled intervention group (PD −6, 95% CI −1, −10) and in the past 3 months from 67% (95% CI 62%, 71%) in the control group to 60% (91% CI 43%, 47%) in the intervention group (PD −7, 95% CI −2, −12). Interventions reduced the prevalence of multiple episodes of antibiotic use in the past 3 months by 7 (95% CI 4, 11) percentage points (control prevalence=25%). On average, interventions reduced the number of days of antibiotic use by 0.93 days (95% CI 0.45, 1.41; control mean=5.2 days) and the number of episodes by 0.18 (95% CI 0.09, 0.26; control mean=1.0 episodes). Overall, effect sizes were similar for individual WSH and nutrition interventions, as well as the WSH and WSH + Nutrition group and the Nutrition and WSH + nutrition group.

### Intervention-mediator effects

Interventions reduced diarrhea prevalence by 33% (95% CI 11%, 49%; control prevalence= 11.6%) and the prevalence of ARI with fever by 26% (95% CI 9%, 40%; control prevalence=13%) (Figure 1A, Figure S4, Table S6). Confidence intervals for effect estimates for ARI, difficulty breathing, and fever alone were less precise or included the null. In the control arm, the prevalence of enteric virus carriage (adenovirus 40/41, norovirus GI, norovirus GII, sapovirus, rotavirus, and astrovirus) was 52% at 14 months, and the mean number of enteric viruses per child was 0.60. Interventions had strong effects on the prevalence of any enteric viral carriage (PR = 0.58, 95% CI 0.49, 0.68) and the prevalence of enteric viral infection with diarrhea symptoms (PR = 0.35, 95% CI 0.24, 0.51). Interventions reduced adenovirus 40/41 prevalence by 40% (95% CI −3%, 65%), norovirus GII prevalence by 33% (95% CI 7%, 52%), and sapovirus prevalence by 49% (95% CI 28%, 64%). Interventions reduced the prevalence of any mediator (diarrhea, ARI, fever, or enteric virus carriage) by 16% (95% CI 8%, 23%). The mean number of enteric viruses was 0.24 viruses (95% CI 0.14, 0.34) lower in intervention vs. control. Effect sizes were similar though slightly weaker for individual nutrition interventions compared to WSH interventions.

**Figure 1:**
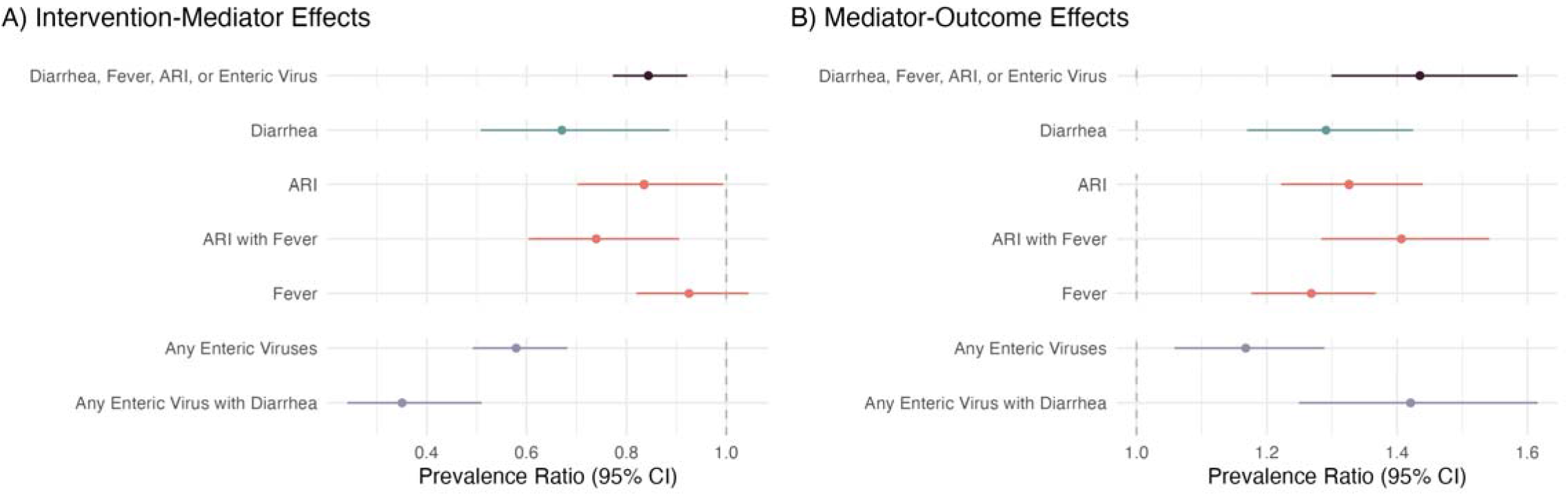
Effects of any WASH and nutrition intervention on mediators, and effects of mediators on caregiver-reported antibiotic use. Prevalence ratios and 95% confidence intervals for (A) effects of any WASH and nutrition intervention on mediators and (B) mediators on any antibiotic use in the past month. Diarrhea, ARI, ARI with Fever, and Fever are all reported by a caregiver under a 7-day lookback period at 14 and 28 months. Any Enteric Virus is the presence of adenovirus 40/41, norovirus GI, norovirus GII, sapovirus, rotavirus, or astrovirus in stool collected at 14 months, and Any Enteric Virus with Diarrhea is the presence of any enteric virus with caregiver reported diarrhea in the prior 7 days at 14 months.

### Mediator-outcome effects

All mediators were associated with a higher prevalence of antibiotic use in the past month with the exception of adenovirus 40/4, norovirus GII, and sapovirus carriage (Figure 1B, Figure S5, Table S7). The strongest associations were for difficulty breathing (PR = 1.47, 95% CI 1.28, 1.70), fever in the past 14 days (PR = 1.48, 95% CI 1.34, 1.63), and enteric virus infection with diarrhea symptoms (PR = 1.42, 95% CI 1.25, 1.62). Diarrhea and ARI were also associated with a higher level of antibiotic use, but associations were slightly weaker. Prevalence of any enteric virus carriage was associated with 17% (95% CI 6%, 29%) higher prevalence antibiotic use and 0.23 (95%. CI −0.38, 0.84) higher mean days of antibiotic use in the past 3 months. Evidence for associations for individual viruses was weak. The prevalence of any mediator (diarrhea, ARI, fever, or enteric virus carriage) was associated with 44% (95% CI 30%, 59%) higher antibiotic use in the past month. Associations were similar but attenuated towards the null for antibiotic use within the past 3 months.

### Mediation

Multiple mediators had indirect effects on antibiotic use in the past month (Figure 2, Figure S6, Table S8). The total effect of interventions on antibiotic use in the past month was −5.5% (−9.9%, −1.2%) percentage points, with reductions of 0.6 percentage points (95% CI 0.1, 1.3; 10.9% of total effect) through reduced diarrhea, 0.7 percentage points (95% CI 0.1, 1.5; 12.7% of total effect) through reduced ARI with fever, 1.8 percentage points (95% CI 0.5, 3.5; 32.7% of total effect) through reduced prevalence of any enteric virus, and 1.1 percentage points (95% CI 0.4, 2.1; 20% of total effect) through reduced prevalence of any enteric virus with diarrhea. Interventions reduced antibiotic use in the past month through any mediator (diarrhea, ARI, fever, or enteric virus carriage) by 2.5 percentage points (95% CI 0.2, 5.3, 45.5% of total effect). Indirect effects on antibiotic use in the past three months were slightly attenuated towards the null. There was only evidence of mediation through diarrhea for WSH interventions and not for nutrition interventions. There was evidence of intervention-mediator interactions in certain analyses; the total NIE was stronger than the pure NIE for any antibiotic use, and we observed the reverse pattern for episodes of antibiotic use (Figure S7, Table S9).

**Figure 2:**
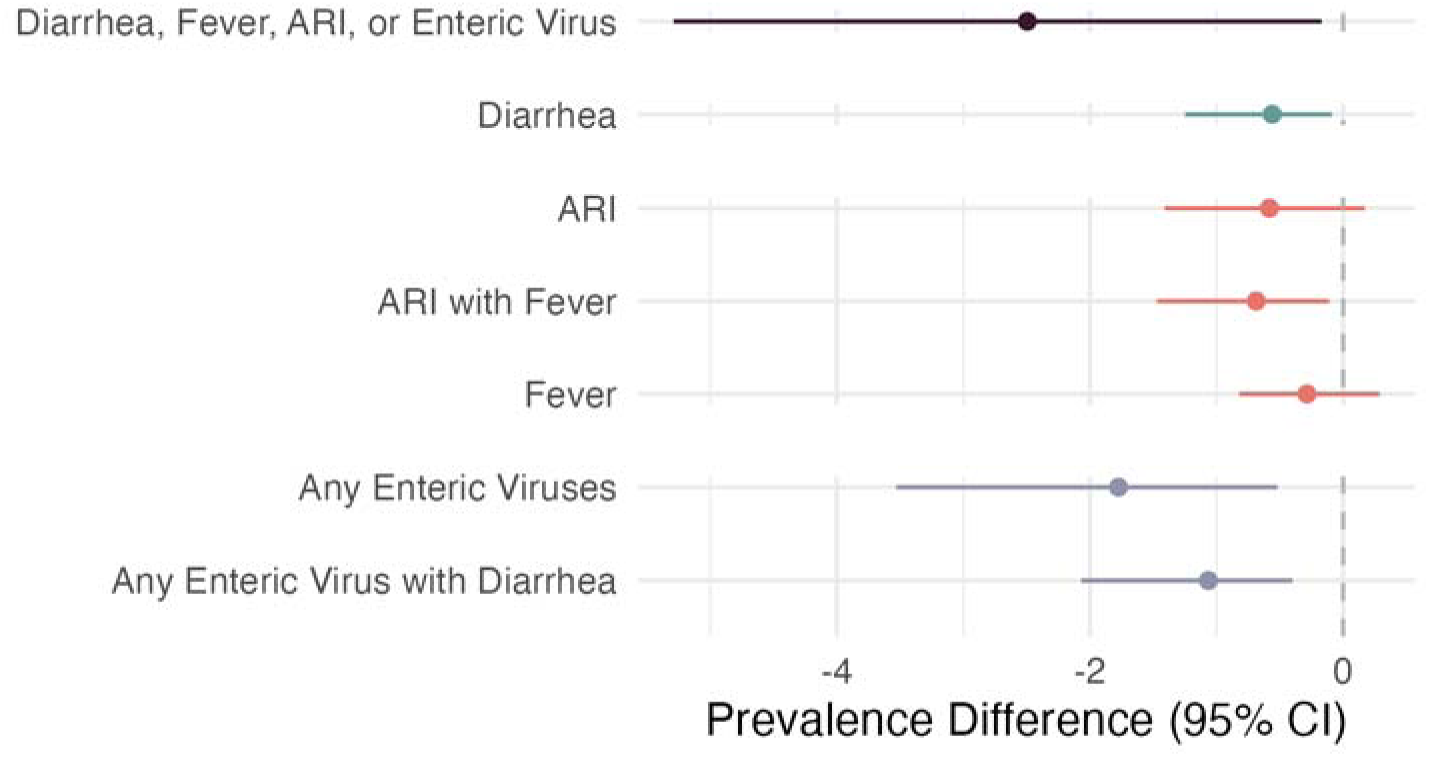
Indirect Effects of WASH and Nutrition interventions on caregiver-reported antibiotic use. Natural indirect effects and 95% confidence intervals of WASH and nutrition interventions on caregiver-reports of any antibiotic use in the past month (total effect = 5.5 percentage points reduction, 95% CI 1.2, 9.9).Diarrhea, ARI, ARI with Fever, and Fever are all reported by a caregiver under a 7-day lookback period at 14 and 28 months. Any Enteric Virus is the presence of adenovirus 40/41, norovirus GI, norovirus GII, sapovirus, rotavirus, or astrovirus in stool collected at 14 months, and Any Enteric Virus with Diarrhea is the presence of any enteric virus with caregiver reported diarrhea in the prior 7 days at 14 months.

### Sensitivity analyses

In analyses using prevalence of enteric viruses with pathogen loads likely to reflect diarrheal etiology, treatment-mediator and mediator-outcome associations were similar (Figure S8-S10, Tables S10-S12), and there was weak evidence of an indirect effect on antibiotic use in the past month (prevalence difference = −0.012 (95% CI −0.030, −0.002)).

### Negative Control

When repeating mediation analyses using bruising prevalence as a negative control for caregiver-reported mediators (e.g., diarrhea, ARI), all 95% confidence intervals for indirect effect estimates contained the null, indicating that mediator misclassification did not have a large impact on our findings (Figure S11, Table S13).

## Discussion

In this mediation analysis of a cluster-randomized trial, effects of low-cost, household-level WASH and nutrition interventions on reduced pediatric antibiotic use occurred via reduced diarrhea and enteric viruses. The strongest mediator was enteric virus carriage, suggesting that interventions effectively reduced antibiotic use by preventing viral enteropathogen infections. Overall, mediation patterns were similar for individual WASH and nutrition interventions, with slightly stronger mediated effects for the WASH and WASH+Nutrition interventions.

About a third of the effect of WASH and nutrition interventions on reduced pediatric antibiotic use was mediated by reduced enteric virus carriage, both with and without diarrhea symptoms. Prior studies have found that viruses are responsible for a large fraction of clinically-attended childhood diarrhea treated with antibiotics in LMICs,^32^ yet diarrhea is frequently treated with antibiotics.^4^ The ratio of appropriately to inappropriately treated diarrhea cases in LMICs is estimated to be 12.6.^4^ These numbers may be larger in community settings where antibiotics are purchased without clinician guidance.

There was weak evidence that interventions reduced antibiotic use by reducing acute respiratory infections with fever. This is likely because WASH and nutrition intervention effects on ARI were relatively modest. While we did not have data on the etiology of ARI in this study, ARI in young children in LMICs are more often of viral etiology than bacterial etiology.^33–36^ Yet, prior studies in Bangladesh and other LMICs have found that 37%-80% of children under 5 with respiratory symptoms were treated with antibiotics.^37–39^

The mediators measured in this study accounted for 45% of the total effect of interventions on antibiotic use. It is possible that there are other important mediators that we did not measure. WASH interventions may reduce skin or eye infections, and such reductions could have resulted in lower antibiotic use. For the nutrition intervention, a key mediating pathway may be improved immune function, however this was beyond the scope of our analysis. A prior analysis found that WASH and nutrition interventions enhanced immunoprotection and immunoregulation and suppressed the immunopathologic response.^40^ Given that nutrition is likely an upstream determinant of immunity and infection, studies have proposed nutrition interventions as a critical tool for reducing AMR in LMICs.^41^ Understanding the immunologic pathways through which nutrition interventions mediate effects on antibiotic use is an important area for future research.

Our findings of reduced antibiotic use do not necessarily reflect changes in antimicrobial resistance carriage. For example, one study in urban Bangladesh found that community-scale water chlorination reduced child antibiotic use by 7% but did not result in reduced prevalence of antimicrobial resistance genes in child stool.^42,43^ While we are not aware of any prior studies that have measured effects of lipid nutrient supplementation on AMR, prior studies have found that dietary diversity was associated with reduced indicators of AMR.^15^ Future studies of WASH and nutrition interventions would benefit from inclusion of antibiotic resistance carriage as an outcome.

Strengths of this study include the use of data from a randomized trial with high intervention uptake and minimized measured and unmeasured confounding of the intervention-mediator relationships. There were several limitations. First, we relied upon caregiver-reported diarrhea, respiratory infection, and antibiotic use, which may be subject to courtesy and recall bias. However, our negative control analysis suggests that there was minimal outcome misclassification. Second, the recall period was 1-3 months for antibiotic use and 7 days for diarrhea and ARI, which were measured concurrently; it is possible that in some cases, antibiotic use preceded diarrhea or ARI. Our results were similar using 1 vs. 3 month recall periods, suggesting that lack of temporal ordering did not have a large influence, but we cannot completely rule it out. Third, we did not consider multiple mediators within a single model; it is possible that our approach did not fully capture complex relationships between mediators. However, joint infections were relatively rare, and we did include an analysis using an indicator for any mediator. Fourth, it is possible that our mediator-outcome models were subject to residual confounding. Finally, our results may not generalize to other populations with differing climatic conditions, pathogen transmission patterns, antibiotic availability, animal ownership, and population density.

In conclusion, our findings shed light on the mechanism through which WASH and nutrition interventions reduce antibiotic use and support a causal interpretation for effects on antibiotic use. Taken together with prior studies, our findings support the use of WASH and nutrition interventions to reduce antimicrobial resistance.

## Data Availability

Data are available online at: https://osf.io/ytmcr

## Acknowledgments

This study was supported by the Gates Foundation (grant OPPGD759 to the University of California, Berkeley). The original trial was implemented by ICDDR,B. Research reported in this publication was supported in part by the National Institutes of Health (NIH) under awards R01HD108196 (PI: Benjamin-Chung), R01AI166671 (PI: Arnold), and F31AI179107 (PI: Nguyen), the Stanford Data Science Scholars Program (Nguyen), and a Stanford University School of Medicine Dean’s Postdoctoral Fellowship (Grembi). The content is solely the responsibility of the authors and does not necessarily represent the official views of the NIH. Jade Benjamin-Chung is a Chan Zuckerberg Biohub Investigator.

## Author contributions

**Conceptualization:** JBC, AE

**Methodology:** JBC, ATN, AE, JG, AL, AM, BFA

**Software:** ATN, GBH

**Validation:** GBH

**Formal analysis:** ATN

**Investigation:** S Ashraf, S Ali, JG, AL, MR

**Data curation:** ZR, S Ali, JG, AL, AE, ATN

**Writing - original draft:** ATN, GBH, JBC

**Writing - reviewing & editing:** All authors

**Visualization:** ATN, GBH

**Supervision:** JBC

**Project administration:** AL, JG, S Ashraf, ZR, S Ali, MR

**Funding acquisition**: BFA

