## Supplement for "Pathways through which water, sanitation, hygiene, and nutrition interventions reduce antibiotic use in young children: a mediation analysis of a cluster-randomized trial": Supplemental Materials.docx

### Supplement 1: Deviations from Pre-Analysis Plan

Prior to this study, we published a pre-analysis plan at <https://osf.io/ytmcr>. Any deviations that we made to this plan are listed below.

1. We pre-specified the inclusion of Norovirus as a mediator. In the trial, both Norovirus GI and GII were measured, but Norovirus GII was the more prevalent. Additionally, the viral load cutoff for diarrheal etiology was validated for Norovirus GII, but not Norovirus GI, in the MAL-ED study. As a result, we chose to focus on Norovirus GII in this analysis, but Norovirus GI is still included in the composite “Any Enteric Virus” mediator.
2. We did not pre-specify the inclusion of the composite mediators that captured (1) diarrhea, ARI, fever, or enteric virus infection at 14 months or (2) diarrhea, ARI, or fever at 14 and 28 months.
3. We estimated the total effect of the intervention on the outcome on the absolute scale using g-computation with logistic models, but these methods were not pre-specified. The addition of these prevalence difference estimates allowed us to present total and mediated effects on the same scale to assist with interpretation.
4. We pre-specified that age would be included as a confounder if it was associated with the outcome, as indicated by a likelihood ratio test p-value < 0.2. However, we chose to adjust all models for age, regardless of the magnitude of the size of the likelihood ratio test p-value, to account for repeated measurements taken for each child at 14 vs 28 months old.
5. We pre-specified the use of log-binomial models when the response variable was categorical, and the use of modified Poisson models only if the log-binomial failed to converge. However, we found that the log-binomial models failed to converge in most cases, so we defaulted to modified Poisson models in all cases in which the response variable was categorical.
6. We pre-specified the inclusion of models with intervention-mediator interactions if the interactions had a large influence on the magnitude of direct and mediated effect estimates. However, we did not pre-specify the criteria that would be used to assess the significance of interaction terms. We chose to include interaction terms if the estimated average causal mediated effect (ACME) was more than 1% different in the intervention vs control groups, or if a t-test to assess if the difference in ACME between arms was not 0 yielded a p-value less than or equal to 0.2.

### Supplement 2. Additional details about study interventions

Interventions were delivered close to the time of index child births. The WSH intervention included chlorine tablets for water treatment and a safe water storage vessel; double-pit latrine upgrades for all latrines in the compound, child potties, and hoes for removing feces; and handwashing stations, which included a soapy water bottle and water for rinsing near kitchens and latrines. The nutrition intervention included promotion of age-appropriate maternal and infant nutrition practices and lipid-based nutrient (LNS) supplements for children from 6-24 months of age. Additional details about the interventions are described elsewhere.^1^ Interventions were given to participants free of charge, and consumables were restocked during trial follow-up. Local promoters visited intervention study compounds to promote intervention uptake weekly in the first 6 months of the trial and once every 2 weeks thereafter. Intervention fidelity was high in the trial as indicated through structured observations and spot checks, where adherence to most interventions exceeded 90%.^2^

1. Luby SP, Rahman M, Arnold BF, et al. Effects of water quality, sanitation, handwashing, and nutritional interventions on diarrhoea and child growth in rural Bangladesh: a cluster randomised controlled trial. Lancet Glob Health 2018; 6: e302–15.

2. Parvez SM, Azad R, Rahman M, et al. Achieving optimal technology and behavioral uptake of single and combined interventions of water, sanitation hygiene and nutrition, in an efficacy trial (WASH benefits) in rural Bangladesh. Trials 2018; 19: 358.

### Supplement 3. Additional details about mediation analysis

To identify potential pathways through which WASH and nutrition interventions affect antibiotic use, we estimated the natural indirect effect (NIE) for mediators with significant mediator-outcome relationships (p-value < 0.05). The NIE is the difference in potential outcomes under the predicted values of the mediator if all children had been in intervention arm versus if all children had been in the control arm. This estimates how WASH interventions modify the prevalence of antibiotic use by shifting the distribution of a given mediator. The NIE can be estimated under potential outcome models that hold intervention status constant to treated (the “total” NIE) or control (the “pure” NIE).^1^ In the absence of intervention -mediator interaction, the total and pure NIEs are equivalent. In the presence of intervention -mediator interaction, the total NIE captures the influence of the interaction, while the pure NIE represents the mediated effect in the absence of interaction. Here, we reported the total NIE for all mediation analyses and additionally reported the pure NIE when we found evidence of intervention -mediator interaction (differences in NIE in the intervention vs control greater than 1% or t-test p-value < 0.2 when an intervention-mediator interaction term was included in the outcome model).

The NIE is estimated while holding either intervention status constant to treated (the “total” NIE) or control (the “pure” NIE). The total NIE captures the influence of the interaction, while the pure NIE represents the mediated effect in the absence of interaction. We considered potential intervention-mediator interaction (differences in NIE in the intervention vs control greater than 1% or t-test p-value < 0.2); in the absence of interaction, the total and pure natural indirect effects are approximately equal, and we reported the total NIE. When interaction was present, we reported the total and pure NIE separately.

1. VanderWeele TJ. A Three-way Decomposition of a Total Effect into Direct, Indirect, and Interactive Effects. Epidemiology. 2013 Mar;24(2):224–32.

13,279 compounds assessed for eligibility

Enrollment

Excluded: 7,728 compounds

7,429 compounds excluded to create buﬀer zones 219 compounds did not meet enrollment criteria 80 compounds declined to participate

720 clusters created and randomly allocated 5,551 compounds randomly allocated

Allocation

**Nutrition + Water + Sanitation + Handwashing (N+WSH)**

90 clusters

686 households

**Control**

180 clusters

1,382 households

**Water + Sanitation**

**+ Handwashing (WSH)**

90 clusters

702 households

**Nutrition**

90 clusters

699 households

**Month 14**

63 clusters

486 children

**Month 28**

66 clusters

514 children

**Month 14**

63 clusters

480 children

**Month 28**

67 clusters

505 children

Subsample Target

**Month 14**

64 clusters

496 children

**Month 28**

66 clusters

514 children

**Month 14**

68 clusters

516 children

**Month 28**

68 clusters

516 children

Follow-up

**Month 14**

74 new children measured

100 children lost to follow- up

9 moved

29 absent

14 withdrew

37 no live birth

11 child death

**Month 28**

25 new children measured

104 children lost to follow- up

28 moved

2 absent

18 withdrew

38 no live birth

18 child death

**Month 14**

78 new children measured 107 children lost to follow- up 17 moved

14 absent

23 withdrew

32 no live birth

21 child death

**Month 28**

28new children measured

142 children lost to follow- up

36 moved

9 absent

42 withdrew

35 no live birth

20 child death

**Month 14**

71 new children measured

101 children lost to follow- up 13 moved

17 absent

17 withdrew

30 no live birth

24 child death

**Month 28**

18 new children measured

115 children lost to follow- up

22 moved

4 absent

30 withdrew

32 no live birth

27 child death

**Month 14**

110 new children measured 140 children lost to follow-up 14 moved

16 absent

62 withdrew

29 no live birth 19 child death

**Month 28**

0 new children measured

158 children lost to follow-up 35 moved

3 absent

72 withdrew

29 no live birth 19 child death

Subsample

Enrollment

**Month 14**

63 clusters

379 children

**Month 28**

67 clusters

401 children

**Month 14**

64 clusters

395 children

**Month 28**

66 clusters

397 children

**Month 14**

63 clusters

377 children

**Month 28**

66 clusters

372 children

**Month 14**

68 clusters

377 children

**Month 28**

68 clusters

358 children

Antibiotic Data

Collection

**Month 14**

63 clusters

348 children

**Month 14**

64 clusters

357 children

**Month 14**

63 clusters

348 children

**Month 14**

67 clusters

312 children

Taqman analysis

**Month 14. Taqman Analyses**

63 clusters

348 children

**Month 14, Other Analyses**

63 clusters

379 children

**Month 28**

67 clusters

401 children

**Month 14. Taqman Analyses**

64 clusters

357 children

**Month 14, Other Analyses**

64 clusters

395 children

**Month 28**

66 clusters

397 children

**Month 14. Taqman Analyses**

63 clusters

348 children

**Month 14, Other Analyses**

63 clusters

377 children

**Month 28**

66 clusters

372 children

**Month 14. Taqman Analyses**

67 clusters

312 children

**Month 14, Other Analyses**

68 clusters

377 children

**Month 28**

68 clusters

358 children

Statistical

Analysis

### Figure S1: Participant flowchart

Flowchart of trial participants that are included in the present analysis, which was restricted to children with non-missing antibiotic use in the follow-up rounds when they were approximately 14 months or 28 months

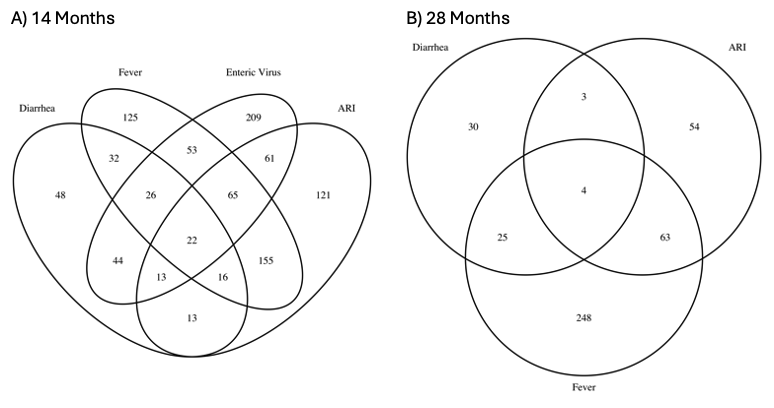

### Figure S2: Joint Prevalence of Mediators

Venn diagram of the number of children with various combinations of co-occurring mediators at measurements taken at approximately 14 and 28 months of age. Diarrhea, ARI, and Fever are all reported by a caregiver under a 7-day lookback period at 14 and 28 months. Enteric Virus is the presence of adenovirus 40/41, norovirus GI, norovirus GII, sapovirus, rotavirus, or astrovirus in stool collected at 14 months.

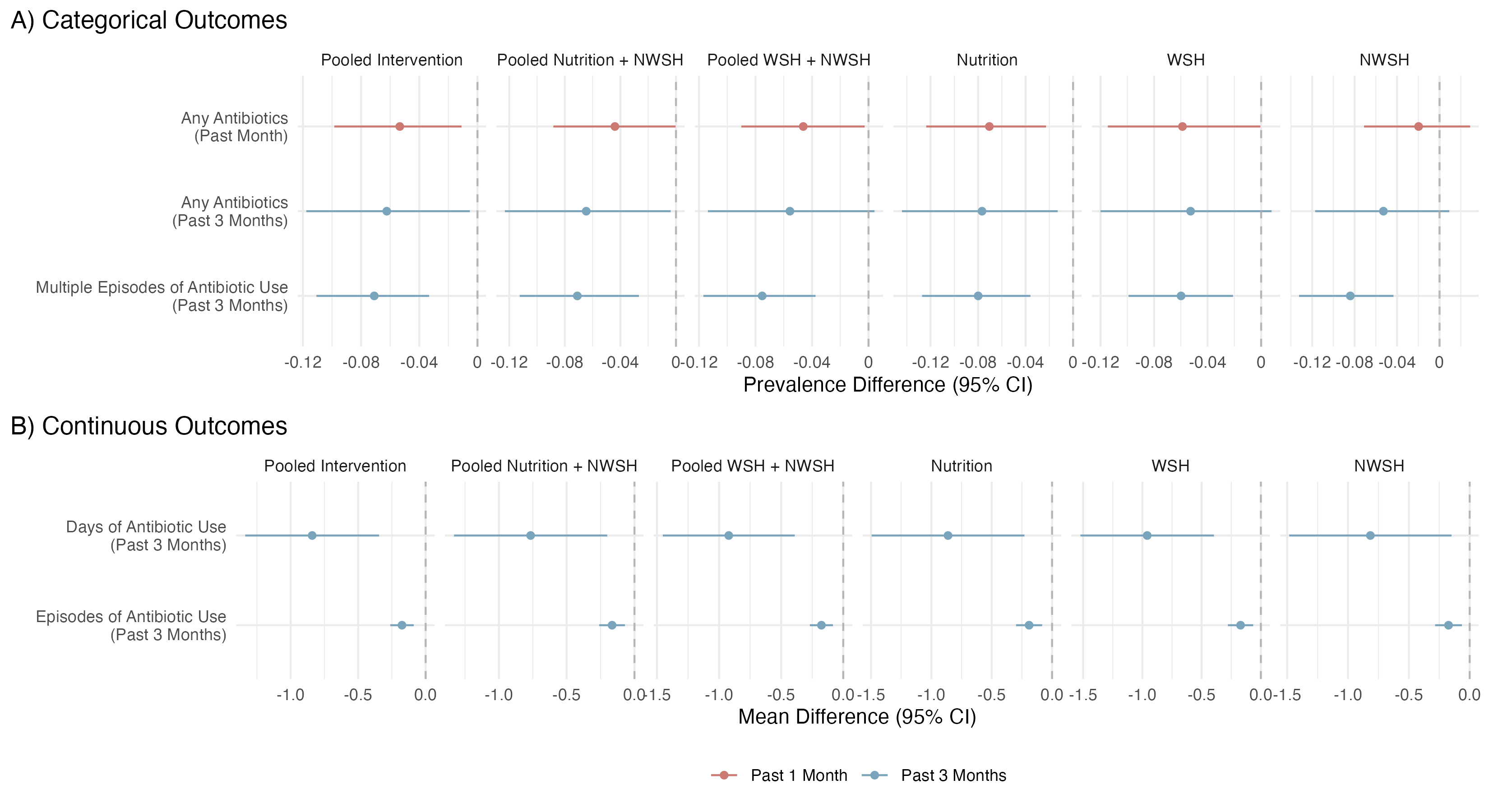

### Figure S3: Intervention-Outcome Effects, All Intervention Groups

(A) Prevalence differences and 95% confidence intervals for the effects of WASH and/or Nutrition interventions on categorical measures of antibiotic use and (B) mean differences and 95% confidence intervals for the effects of WASH and/or Nutrition interventions on continuous measures of antibiotic use. Antibiotic use is reported by a caregiver under either a 1- or 3-month look back period at 14 and 28 months.

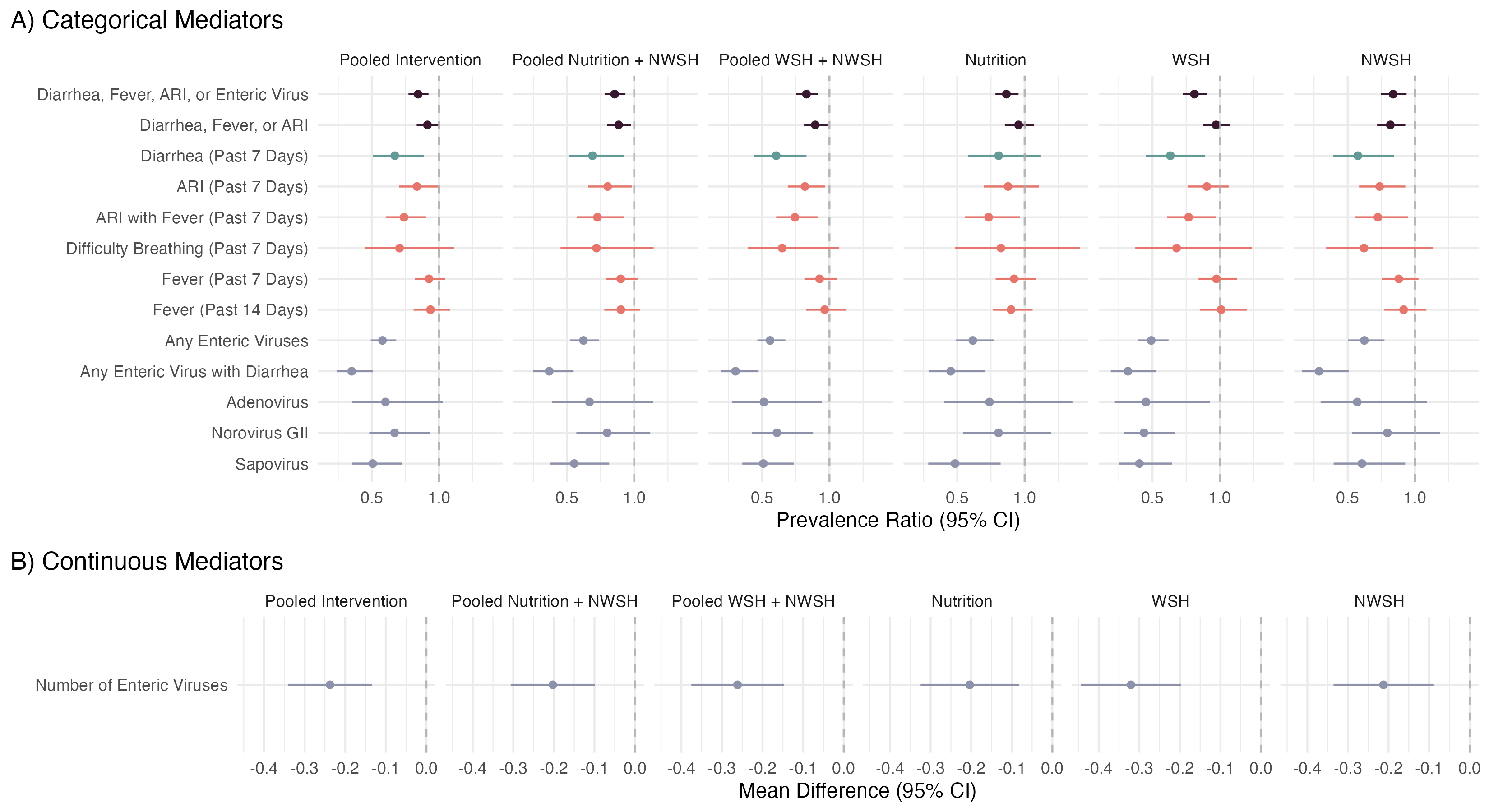

### Figure S4: Intervention-Mediator Effects, All Intervention Groups and Mediators

(A) Prevalence differences and 95% confidence intervals for the effects of WASH and/or Nutrition interventions on categorical mediators and (B) Prevalence differences and 95% confidence intervals for the effects of WASH and/or Nutrition interventions on continuous mediators. Diarrhea, ARI, ARI with Fever, and Fever are all reported by a caregiver under a 7-day lookback period at 14 and 28 months. Any Enteric Virus is the presence of adenovirus 40/41, norovirus GI, norovirus GII, sapovirus, rotavirus, or astrovirus in stool collected at 14 months, and Any Enteric Virus with Diarrhea is the presence of any enteric virus with caregiver reported diarrhea in the prior 7 days at 14 months.

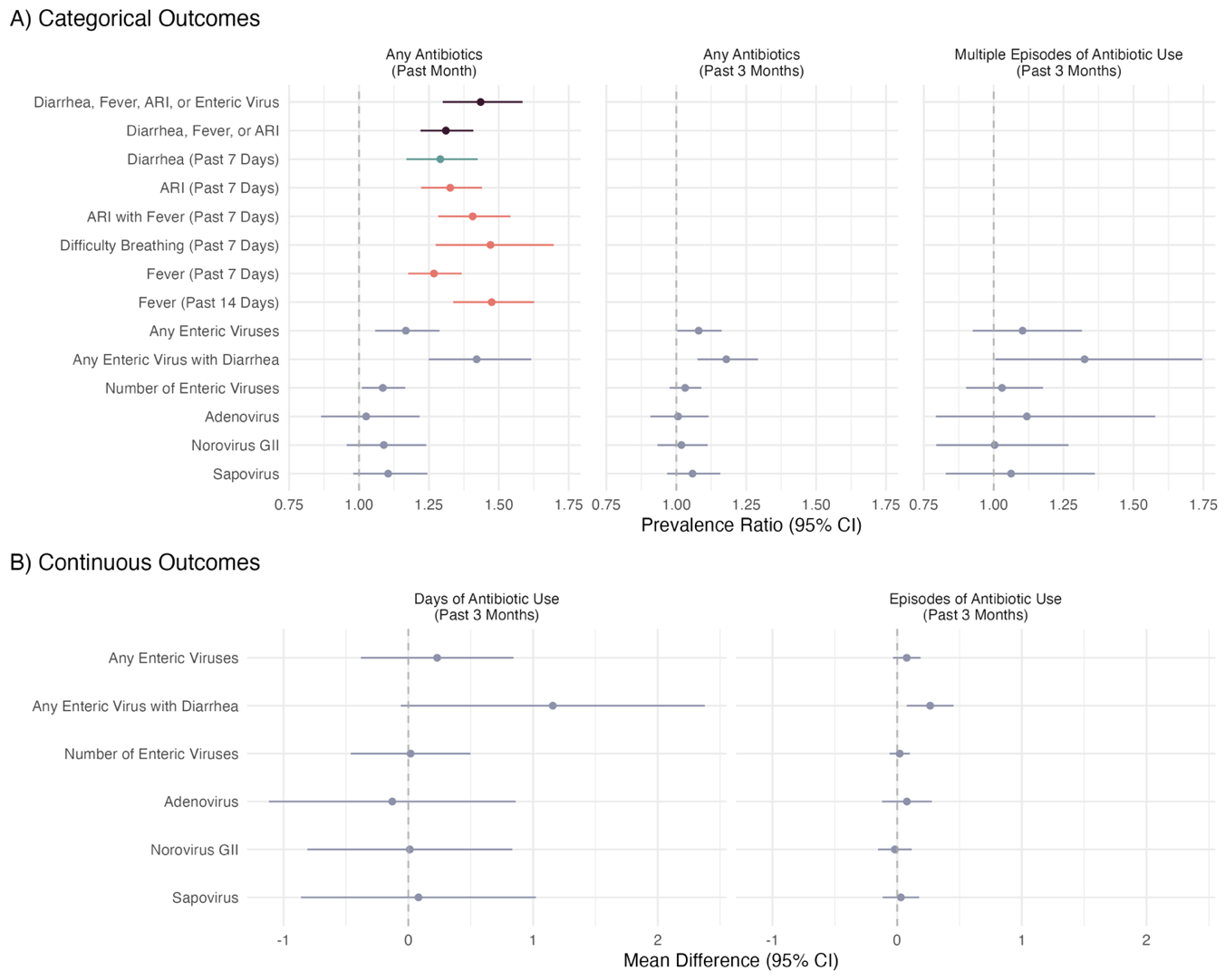

### Figure S5: Mediator-Outcome Effects, All Intervention Groups and Mediators

(A) Prevalence ratios and 95% confidence intervals for the effect of potential mediators on categorical measures of antibiotic use (B) Mean differences and 95% confidence intervals for the effect of potential mediators on continuous measures of antibiotic use. Diarrhea, ARI, ARI with Fever, and Fever are all reported by a caregiver under a 7-day lookback period at 14 and 28 months. Any Enteric Virus is the presence of adenovirus 40/41, norovirus GI, norovirus GII, sapovirus, rotavirus, or astrovirus in stool collected at 14 months, and Any Enteric Virus with Diarrhea is the presence of any enteric virus with caregiver reported diarrhea in the prior 7 days at 14 months. Antibiotic use is reported by a caregiver under either a 1- or 3-month lookback period at 14 and 28 months.

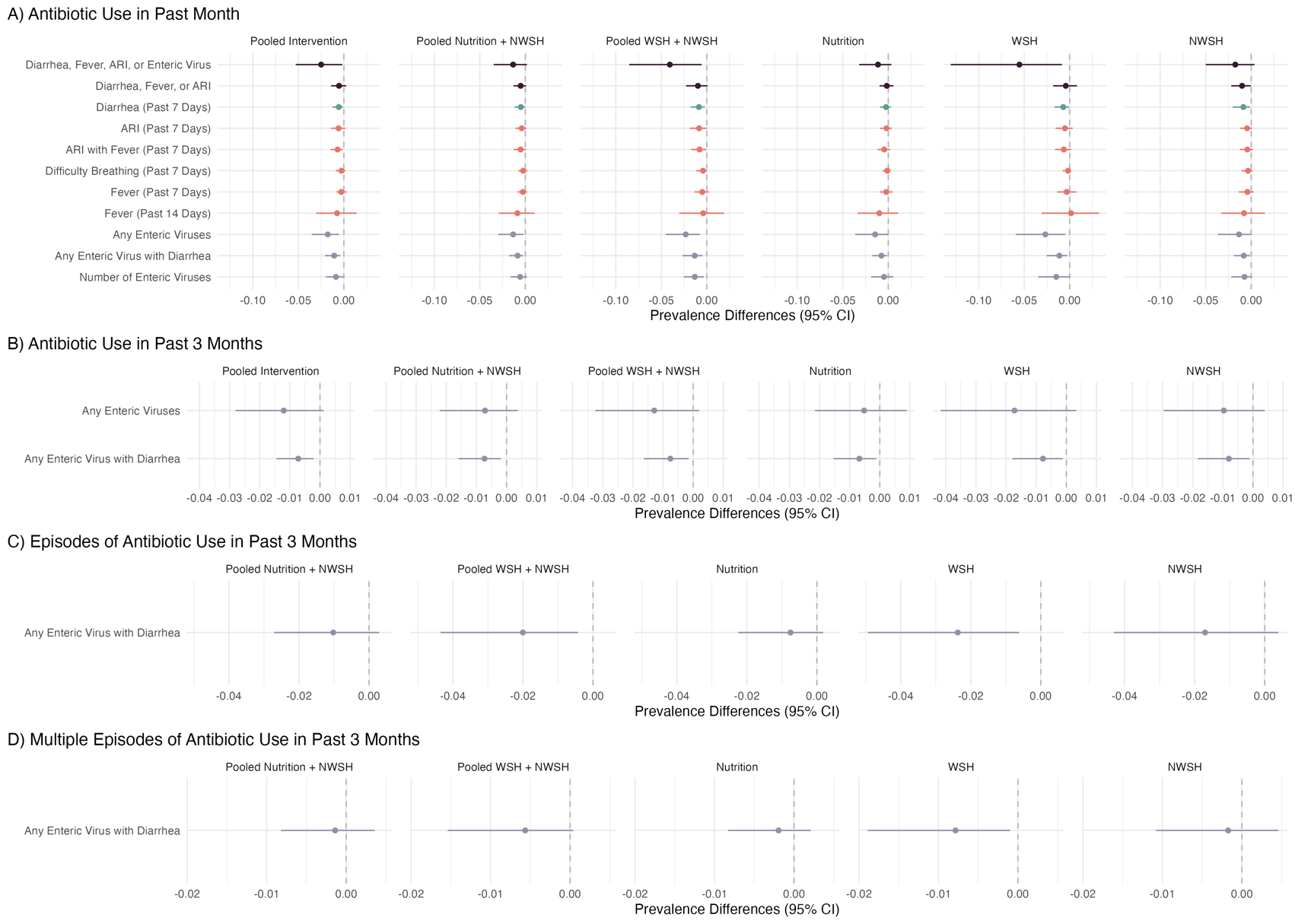

### Figure S6: Mediated Effects, All Intervention Groups and Mediators

Total natural indirect effect estimates and 95% confidence intervals of WASH and/or Nutrition interventions on antibiotic use, for significant mediator-outcome relationships (p-val < 0.05). Diarrhea, ARI, ARI with Fever, and Fever are all reported by a caregiver under a 7-day lookback period at 14 and 28 months. Any Enteric Virus is the presence of adenovirus 40/41, norovirus GI, norovirus GII, sapovirus, rotavirus, or astrovirus in stool collected at 14 months, and Any Enteric Virus with Diarrhea is the presence of any enteric virus with caregiver reported diarrhea in the prior 7 days at 14 months. Antibiotic use is reported by a caregiver under either a 1- or 3-month lookback period at 14 and 28 months.

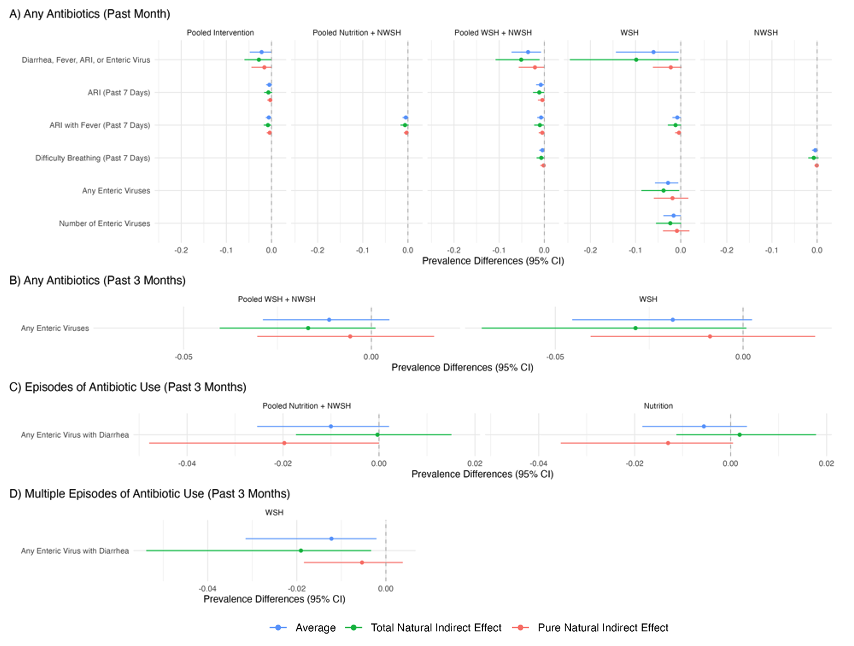

### Figure S7: Mediated Effects, with Significant Intervention-Mediator Interactions

Estimates and 95% CIs of total, pure, and averaged natural indirect effects, computed only when the difference in total and pure natural indirect effects was greater than 1% or had a t-test p-value < 0.2 when outcome models included an intervention-mediator interaction term.

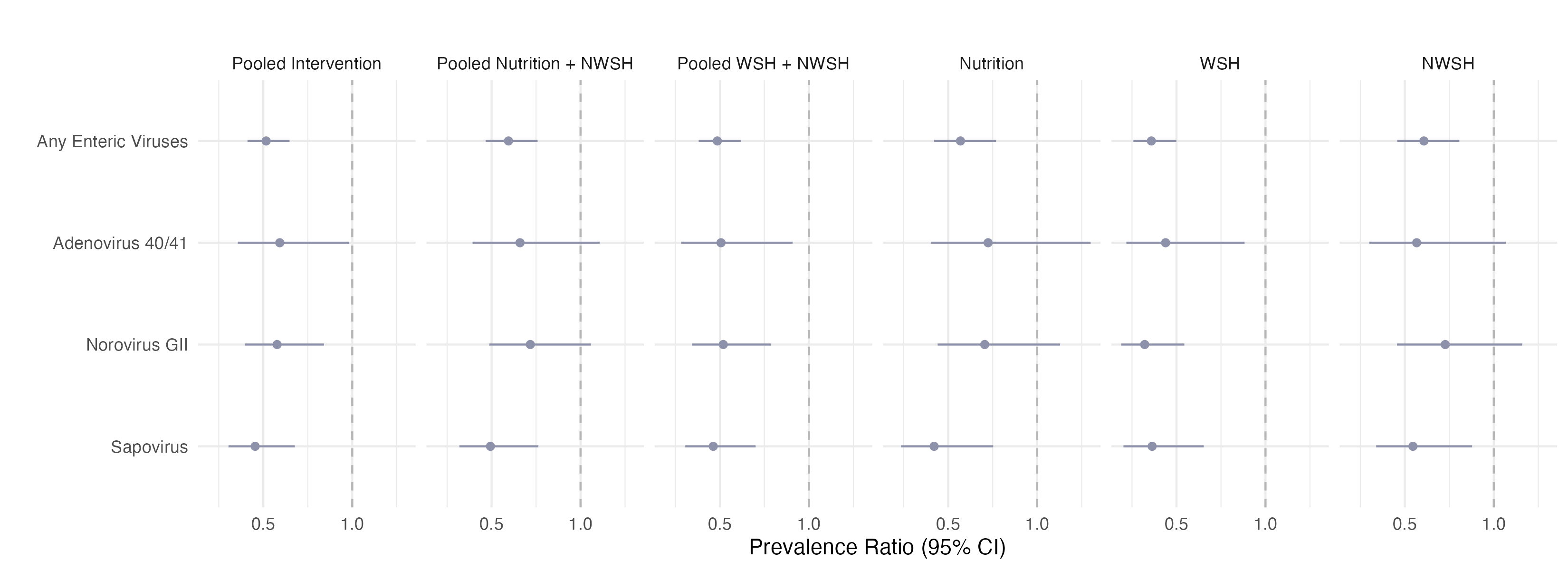

### Figure S8: Intervention-Mediator Effects, for Enteric Viruses with Diarrheal Etiology

Prevalence ratios and 95% confidence intervals for the effects of WASH and/or Nutrition interventions for enteric virus carriage at 14 months with pathogen loads that reflect diarrheal etiology based on published Ct cutoff values from the MAL-ED study. Any Enteric Virus is the presence of Adenovirus 40/41, Norovirus GII, or Sapovirus that exceeded the etiology cutoff.

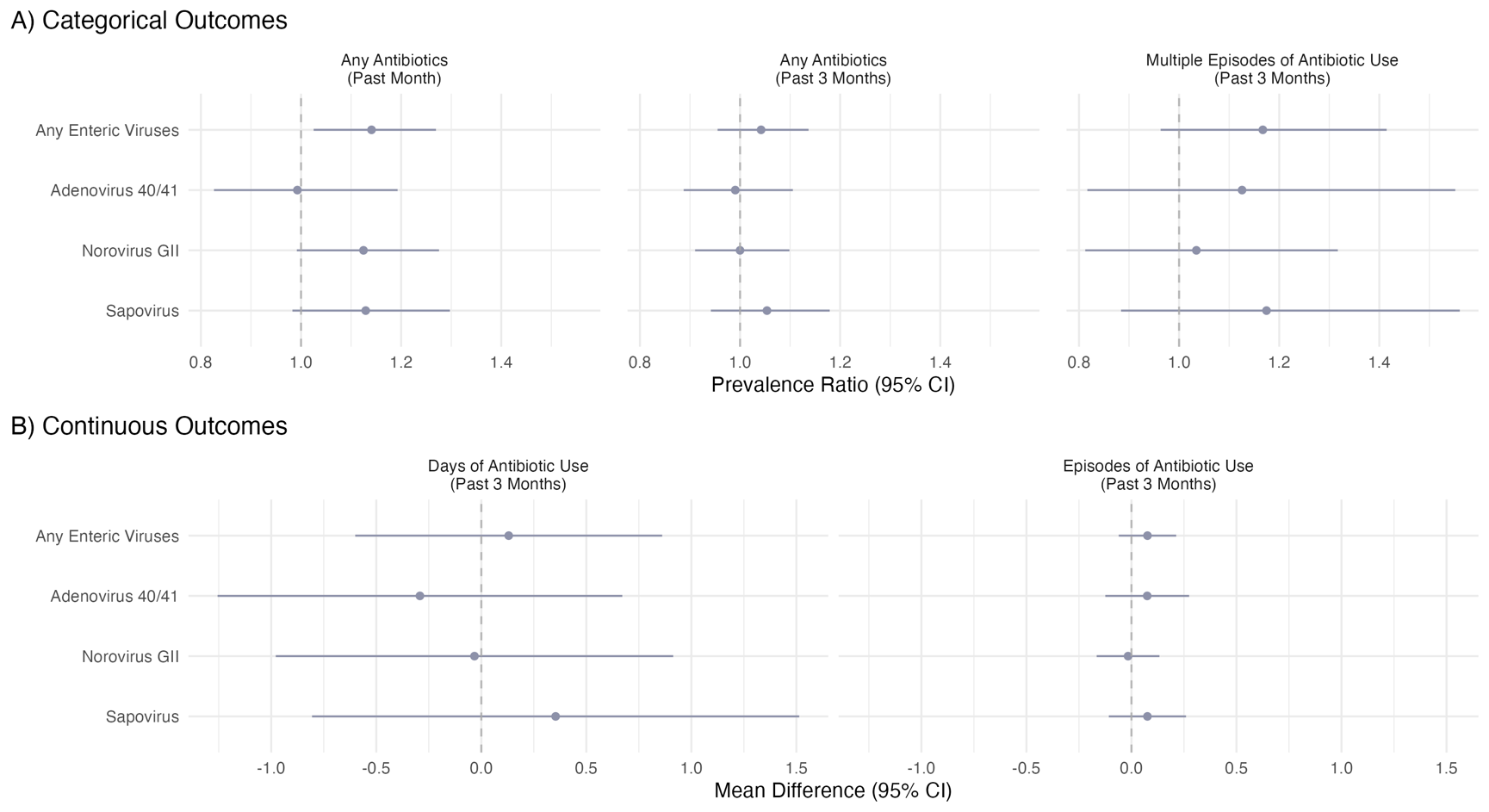

### Figure S9: Mediator-Outcome Effects, for Enteric Viruses with Diarrheal Etiology

(A) Prevalence ratios and 95% confidence intervals for the effect of enteric virus carriage at 14 months with pathogen loads that reflect diarrheal etiology on categorical measures of antibiotic use and (B) Mean differences and 95% confidence intervals for the effect of enteric virus carriage at 14 months with pathogen loads that reflect diarrheal etiology on continuous measures of antibiotic use. Diarrheal etiology is assessed using published Ct cutoff values from the MAL-ED study. Any Enteric Virus is the presence of Adenovirus 40/41, Norovirus GII, or Sapovirus that exceeded the etiology cutoff. Antibiotic use is reported by a caregiver under either a 1- or 3-month lookback period at 14 and 28 months.

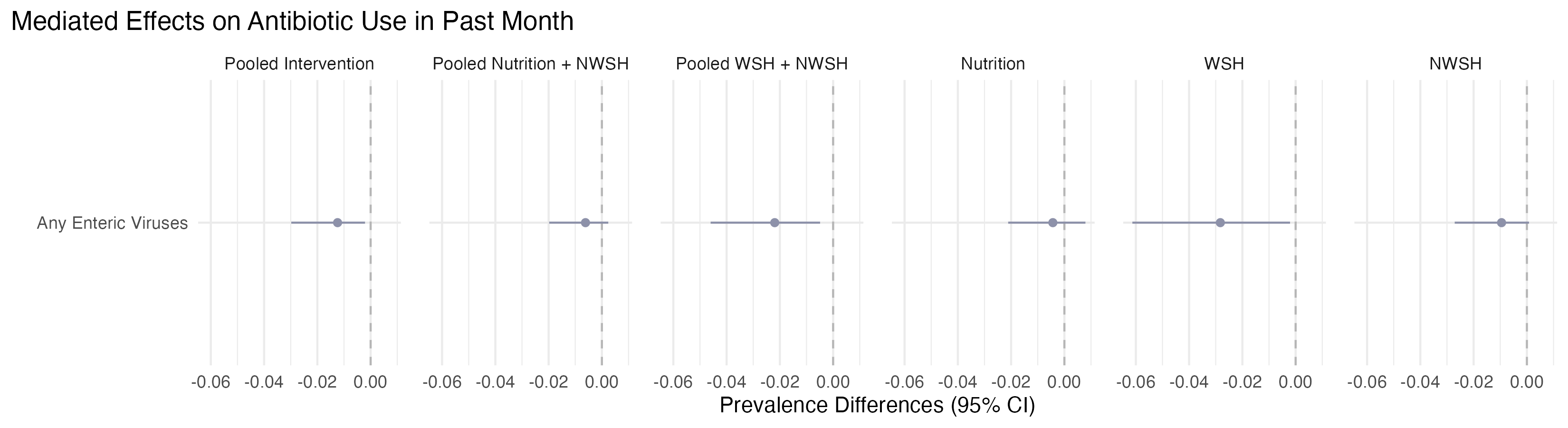

### Figure S10: Mediated Effects, for Enteric Viruses with Diarrheal Etiology

Total natural indirect effect estimates and 95% confidence intervals of WASH and/or Nutrition interventions on antibiotic use, for significant mediator-outcome relationships (p-val < 0.05) involving enteric virus carriage at 14 months with pathogen loads that reflect diarrheal etiology. Diarrheal etiology is assessed using published Ct cutoff values from the MAL-ED study. Any Enteric Virus is the presence of Adenovirus 40/41, Norovirus GII, or Sapovirus that exceeded the etiology cutoff. Antibiotic use is reported by a caregiver under either a 1- or 3-month lookback period at 14 and 28 months.

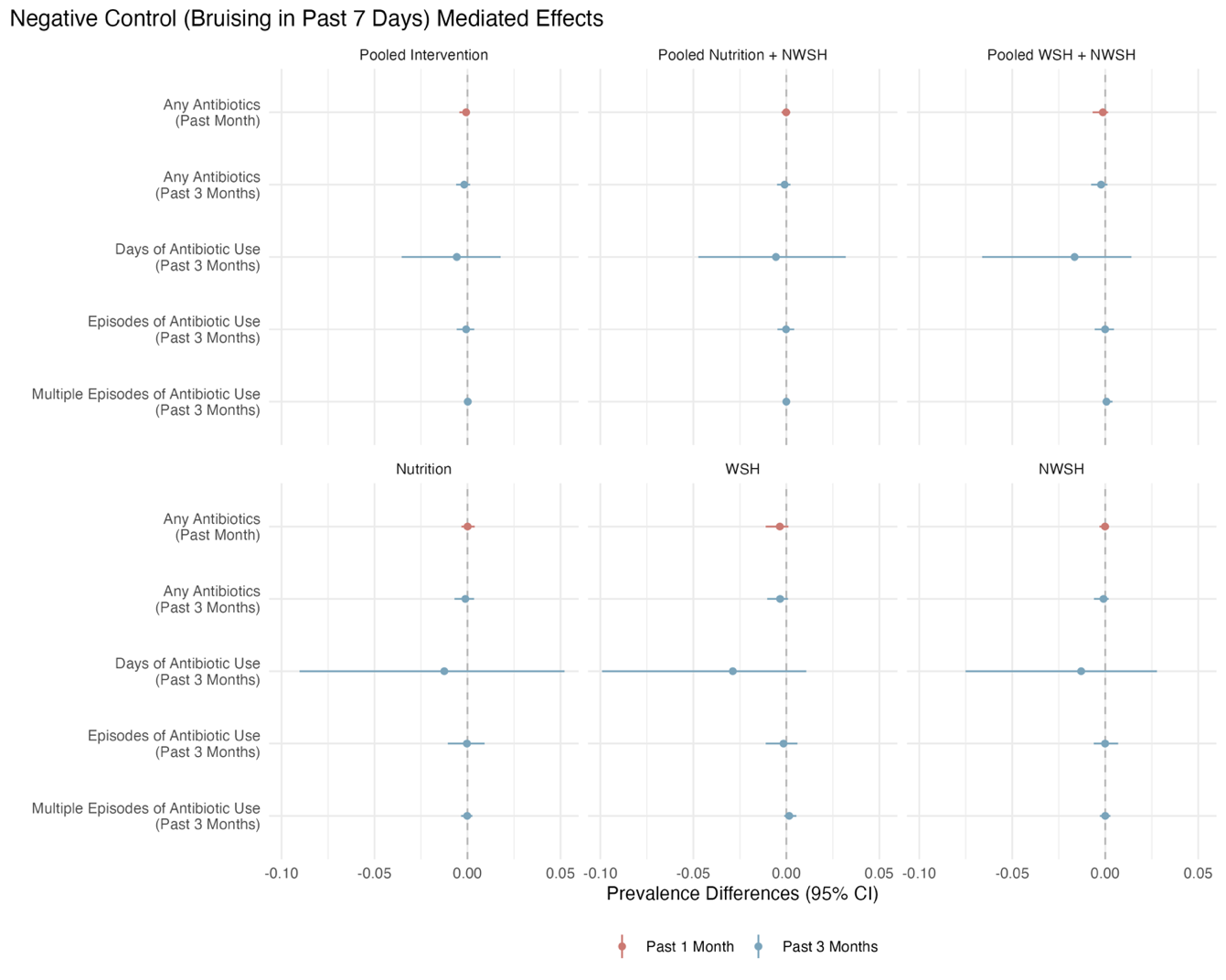

### Figure S11: Mediated Effects, Negative Control

Total natural indirect effect estimates and 95% confidence intervals of WASH and/or Nutrition interventions on antibiotic use through pathways involving bruising, a negative control. Bruising is reported by a caregiver under a 7-day lookback period at 14 and 28 months. Antibiotic use is reported by a caregiver under either a 1- or 3-month lookback period at 14 and 28 months.

### Table S1: Covariate Adjustments in Mediation Models

| **Intervention Group** | **Mediator** | **Outcome** | **Adjusted Covariates** |
| --- | --- | --- | --- |
| Pooled Intervention | Diarrhea, Fever, ARI, or Enteric Virus | Any antibiotic use in past month | Measurement date, Child's sex |
|  | Diarrhea, Fever, or ARI | Any antibiotic use in past month | Child's sex, Household Wall Material |
|  | Diarrhea | Any antibiotic use in past month | Child's sex, Household Wall Material |
|  | ARI | Any antibiotic use in past month | Child's sex, Household Wall Material |
|  | ARI with Fever | Any antibiotic use in past month | Child's sex, Household Wall Material |
|  | Difficulty Breathing | Any antibiotic use in past month | Child's sex, Household Wall Material |
|  | Fever (Past 7 Days) | Any antibiotic use in past month | Child's sex, Household Wall Material |
|  | Fever (Past 14 Days) | Any antibiotic use in past month | Measurement date, Child's sex, Distance to water source |
|  | Any Enteric Virus | Any antibiotic use in past month | Measurement date, Child's sex |
|  |  | Any antibiotic use in past 3 months | Child's sex |
|  | Any Enteric Virus with Diarrhea | Any antibiotic use in past month | Child's sex, Household Wall Material |
|  |  | Any antibiotic use in past 3 months | Child's sex, Mother's height, Household Wealth Index |
|  |  | Episodes of antibiotic use in past 3 months | Child's sex, Mother's height, Mother's education, Number of individuals in household under age 18, Household Wall Material, Household Wealth Index |
|  |  | Multiple episodes of antibiotic use | Measurement date, Child's sex, Mother's height, Mother's education, Number of individuals in household under age 18 |
|  | Number of Enteric Viruses | Any antibiotic use in past month | Measurement date, Child's sex |
| Pooled Nutrition and Nutrition + WSH | Diarrhea, Fever, ARI, or Enteric Virus | Any antibiotic use in past month | Measurement date, Child's sex |
|  | Diarrhea, Fever, or ARI | Any antibiotic use in past month | Measurement date, Child's sex |
|  | Diarrhea | Any antibiotic use in past month | Measurement date, Child's sex |
|  | ARI | Any antibiotic use in past month | Measurement date, Child's sex |
|  | ARI with Fever | Any antibiotic use in past month | Measurement date, Child's sex |
|  | Difficulty Breathing | Any antibiotic use in past month | Measurement date, Child's sex |
|  | Fever (Past 7 Days) | Any antibiotic use in past month | Measurement date, Child's sex |
|  | Fever (Past 14 Days) | Any antibiotic use in past month | Measurement date, Child's sex |
|  | Any Enteric Virus | Any antibiotic use in past month | Measurement date, Child's sex |
|  |  | Any antibiotic use in past 3 months | Measurement date, Child's sex |
|  | Any Enteric Virus with Diarrhea | Any antibiotic use in past month | Measurement date, Child's sex |
|  |  | Any antibiotic use in past 3 months | Child's sex, Mother's height, Household Wealth Index |
|  |  | Episodes of antibiotic use in past 3 months | Measurement date, Child's sex, Mother's education, Number of individuals in household under age 18, Household Wealth Index |
|  |  | Multiple episodes of antibiotic use | Measurement date, Child's sex, Mother's education, Number of individuals in household under age 18 |
|  | Number of Enteric Viruses | Any antibiotic use in past month | Measurement date, Child's sex |
| Pooled WSH and Nutrition + WSH | Diarrhea, Fever, ARI, or Enteric Virus | Any antibiotic use in past month | Child's sex, Household Wall Material |
|  | Diarrhea, Fever, or ARI | Any antibiotic use in past month | Child's sex, Household Wall Material, Household Wealth Index |
|  | Diarrhea | Any antibiotic use in past month | Child's sex, Household Wall Material, Household Wealth Index |
|  | ARI | Any antibiotic use in past month | Child's sex, Household Wall Material, Household Wealth Index |
|  | ARI with Fever | Any antibiotic use in past month | Child's sex, Household Wall Material, Household Wealth Index |
|  | Difficulty Breathing | Any antibiotic use in past month | Child's sex, Household Wall Material, Household Wealth Index |
|  | Fever (Past 7 Days) | Any antibiotic use in past month | Child's sex, Household Wall Material, Household Wealth Index |
|  | Fever (Past 14 Days) | Any antibiotic use in past month | Child's sex, Distance to water source, Household Wall Material |
|  | Any Enteric Virus | Any antibiotic use in past month | Child's sex, Household Wall Material |
|  |  | Any antibiotic use in past 3 months | Number of individuals in compound |
|  | Any Enteric Virus with Diarrhea | Any antibiotic use in past month | Child's sex, Household Wall Material, Household Wealth Index |
|  |  | Any antibiotic use in past 3 months | Child's sex, Household Wealth Index |
|  |  | Episodes of antibiotic use in past 3 months | Child's sex, Household Wealth Index |
|  |  | Multiple episodes of antibiotic use | Child's sex, Number of individuals in household under age 18 |
|  | Number of Enteric Viruses | Any antibiotic use in past month | Child's sex, Household Wall Material |
| Nutrition | Diarrhea, Fever, ARI, or Enteric Virus | Any antibiotic use in past month | Measurement date, Number of individuals in compound |
|  | Diarrhea, Fever, or ARI | Any antibiotic use in past month | Measurement date, Child's sex, Mother's age |
|  | Diarrhea | Any antibiotic use in past month | Measurement date, Child's sex, Mother's age |
|  | ARI | Any antibiotic use in past month | Measurement date, Child's sex, Mother's age |
|  | ARI with Fever | Any antibiotic use in past month | Measurement date, Child's sex, Mother's age |
|  | Difficulty Breathing | Any antibiotic use in past month | Measurement date, Child's sex, Mother's age |
|  | Fever (Past 7 Days) | Any antibiotic use in past month | Measurement date, Child's sex, Mother's age |
|  | Fever (Past 14 Days) | Any antibiotic use in past month | Measurement date, Number of individuals in compound |
|  | Any Enteric Virus | Any antibiotic use in past month | Measurement date, Number of individuals in compound |
|  |  | Any antibiotic use in past 3 months | Measurement date |
|  | Any Enteric Virus with Diarrhea | Any antibiotic use in past month | Measurement date, Birth order, Mother's age, Household Floor Material |
|  |  | Any antibiotic use in past 3 months | Child's sex, Mother's age, Household Floor Material, Household Wealth Index |
|  |  | Episodes of antibiotic use in past 3 months | Measurement date, Child's sex, Mother's age, Mother's education, Household Floor Material |
|  |  | Multiple episodes of antibiotic use | Measurement date, Mother's education, Number of individuals in household under age 18, Household Floor Material |
|  | Number of Enteric Viruses | Any antibiotic use in past month | Measurement date, Number of individuals in compound |
| WSH | Diarrhea, Fever, ARI, or Enteric Virus | Any antibiotic use in past month | Household Wall Material |
|  | Diarrhea, Fever, or ARI | Any antibiotic use in past month | Household Wealth Index |
|  | Diarrhea | Any antibiotic use in past month | Household Wealth Index |
|  | ARI | Any antibiotic use in past month | Household Wealth Index |
|  | ARI with Fever | Any antibiotic use in past month | Household Wealth Index |
|  | Difficulty Breathing | Any antibiotic use in past month | Household Wealth Index |
|  | Fever (Past 7 Days) | Any antibiotic use in past month | Household Wealth Index |
|  | Fever (Past 14 Days) | Any antibiotic use in past month |  |
|  | Any Enteric Virus | Any antibiotic use in past month | Household Wall Material |
|  |  | Any antibiotic use in past 3 months |  |
|  | Any Enteric Virus with Diarrhea | Any antibiotic use in past month | Mother's age, Household Wealth Index |
|  |  | Any antibiotic use in past 3 months | Household Wealth Index |
|  |  | Episodes of antibiotic use in past 3 months | Child's sex, Household Wealth Index |
|  |  | Multiple episodes of antibiotic use | Child's sex, Number of individuals in household under age 18 |
|  | Number of Enteric Viruses | Any antibiotic use in past month | Household Wall Material |
| Nutrition + WSH | Diarrhea, Fever, ARI, or Enteric Virus | Any antibiotic use in past month | Measurement date, Child's sex, Distance to water source, Household Wall Material |
|  | Diarrhea, Fever, or ARI | Any antibiotic use in past month | Measurement date, Child's sex |
|  | Diarrhea | Any antibiotic use in past month | Measurement date, Child's sex |
|  | ARI | Any antibiotic use in past month | Measurement date, Child's sex |
|  | ARI with Fever | Any antibiotic use in past month | Measurement date, Child's sex |
|  | Difficulty Breathing | Any antibiotic use in past month | Measurement date, Child's sex |
|  | Fever (Past 7 Days) | Any antibiotic use in past month | Measurement date, Child's sex |
|  | Fever (Past 14 Days) | Any antibiotic use in past month | Measurement date, Child's sex |
|  | Any Enteric Virus | Any antibiotic use in past month | Measurement date, Child's sex, Distance to water source, Household Wall Material |
|  |  | Any antibiotic use in past 3 months | Child's sex |
|  | Any Enteric Virus with Diarrhea | Any antibiotic use in past month | Measurement date, Child's sex |
|  |  | Any antibiotic use in past 3 months | Child's sex, Household Wealth Index |
|  |  | Episodes of antibiotic use in past 3 months | Child's sex, Number of individuals in household under age 18, Household Wealth Index |
|  |  | Multiple episodes of antibiotic use | Child's sex, Number of individuals in household under age 18 |
|  | Number of Enteric Viruses | Any antibiotic use in past month | Measurement date, Child's sex, Distance to water source, Household Wall Material |

Covariates used for adjustment in intervention-mediator and mediator-outcome models for mediation analyses. We adjusted mediator-outcome models for any of the potential confounders with a likelihood ratio test p-value < 0.2: time of fecal sample/data collection (measured in 3-month periods); sex, and birth order; mother’s age, height and education; household food insecurity; number of individuals <18 years in household; number of individuals living in compound; distance to household’s primary drinking water source; housing materials; and a household wealth index calculated from the first principal component of a principal components analysis of household assets. All covariates were measured at baseline except for month of measurement, child age, and child sex.

### Table S2: Distribution of Baseline Characteristics, Mediators, and Antibiotic Use Outcomes, All Intervention Groups

|  | **Nutrition**  **(N = 419)** | **WSH**  **(N= 434)** | **NWSH**  **(N = 441)** | **Nutrition + NWSH**  **(N = 860)** | **WSH + NWSH**  **(N = 875)** | **Control**  **(N = 422)** |
| --- | --- | --- | --- | --- | --- | --- |
| **BASELINE COVARIATES** | | | | | | |
| Month of Measurement | | | | | | |
| *Feb-May* | 236 (31.51%) | 246 (31.06%) | 207 (26.54%) | 443 (28.97%) | 453 (28.82%) | 253 (34.42%) |
| *Jun-Sep* | 270 (36.05%) | 329 (41.54%) | 327 (41.92%) | 597 (39.05%) | 656 (41.73%) | 258 (35.1%) |
| *Oct-Jan* | 243 (32.44%) | 217 (27.4%) | 246 (31.54%) | 489 (31.98%) | 463 (29.45%) | 224 (30.48%) |
| Child's age | 15.71 (7.43) | 15.68 (7.44) | 15.95 (7.37) | 15.83 (7.39) | 15.81 (7.40) | 16.43 (7.40) |
| Child's sex | | | | | | |
| *Female* | 361 (48.20%) | 371 (46.84%) | 410 (52.56%) | 771 (50.43%) | 781 (49.68%) | 372 (50.61%) |
| *Male* | 388 (51.80%) | 421 (53.16%) | 370 (47.44%) | 758 (49.57%) | 791 (50.32%) | 363 (49.39%) |
| Birth Order | | | | | | |
| *1* | 269 (35.91%) | 248 (31.31%) | 269 (34.49%) | 538 (35.19%) | 517 (32.89%) | 272 (37.01%) |
| *2+* | 463 (61.82%) | 529 (66.79%) | 494 (63.33%) | 957 (62.59%) | 1023 (65.08%) | 416 (56.6%) |
| *Missing* | 17 (2.27%) | 15 (1.89%) | 17 (2.18%) | 34 (2.22%) | 32 (2.04%) | 47 (6.39%) |
| Mother's age | 23.75 (5.00) | 24.46 (5.38) | 24.24 (5.48) | 24.00 (5.26) | 24.35 (5.43) | 23.45 (4.85) |
| Mother's height | 150.41 (5.55) | 150.44 (5.46) | 150.05 (5.22) | 150.22 (5.38) | 150.25 (5.34) | 150.84 (5.16) |
| Mother's education | | | | | | |
| *No education* | 112 (14.95%) | 96 (12.12%) | 126 (16.15%) | 238 (15.57%) | 222 (14.12%) | 78 (10.61%) |
| *Primary (1-5y)* | 220 (29.37%) | 222 (28.03%) | 250 (32.05%) | 470 (30.74%) | 472 (30.03%) | 179 (24.35%) |
| *Secondary (>5y)* | 417 (55.67%) | 474 (59.85%) | 404 (51.79%) | 821 (53.7%) | 878 (55.85%) | 478 (65.03%) |
| Household food insecurity | | | | | | |
| *Food Secure* | 538 (71.83%) | 534 (67.42%) | 554 (71.03%) | 1092 (71.42%) | 1088 (69.21%) | 543 (73.88%) |
| *Mildly Food Insecure* | 57 (7.61%) | 77 (9.72%) | 67 (8.59%) | 124 (8.11%) | 144 (9.16%) | 62 (8.44%) |
| *Moderately/Severely*  *Food Insecure* | 154 (20.56%) | 181 (22.85%) | 159 (20.38%) | 313 (20.47%) | 340 (21.63%) | 130 (17.69%) |
| Number of individuals in  household under age 18 | 1.52 (1.30) | 1.65 (1.21) | 1.72 (1.29) | 1.62 (1.30) | 1.68 (1.25) | 1.56 (1.25) |
| Number of individuals in  compound | 11.33 (6.46) | 11.43 (6.38) | 11.46 (6.82) | 11.40 (6.65) | 11.44 (6.60) | 10.10 (6.07) |
| Distance to water source | 0.76 (1.55) | 1.14 (6.19) | 0.76 (2.23) | 0.76 (1.93) | 0.95 (4.67) | 0.75 (1.53) |
| Household Roof Material | 738 (98.53%) | 781 (98.61%) | 770 (98.72%) | 1508 (98.63%) | 1551 (98.66%) | 729 (99.18%) |
| Household Wall Material | 507 (67.69%) | 498 (62.88%) | 525 (67.31%) | 1032 (67.5%) | 1023 (65.08%) | 449 (61.09%) |
| Household Floor Material | 89 (11.88%) | 91 (11.49%) | 94 (12.05%) | 183 (11.97%) | 185 (11.77%) | 121 (16.46%) |
| Household Wealth Index | | | | | | |
| *Wealth Q1* | 176 (23.5%) | 180 (22.73%) | 160 (20.51%) | 336 (21.98%) | 340 (21.63%) | 100 (13.61%) |
| *Wealth Q2* | 178 (23.77%) | 167 (21.09%) | 195 (25%) | 373 (24.4%) | 362 (23.03%) | 138 (18.78%) |
| *Wealth Q3* | 130 (17.36%) | 186 (23.48%) | 156 (20%) | 286 (18.71%) | 342 (21.76%) | 185 (25.17%) |
| *Wealth Q4* | 178 (23.77%) | 173 (21.84%) | 172 (22.05%) | 350 (22.89%) | 345 (21.95%) | 193 (26.26%) |
| *Missing* | 87 (11.62%) | 86 (10.86%) | 97 (12.44%) | 184 (12.03%) | 183 (11.64%) | 119 (16.19%) |
| **MEDIATORS** | | | | | | |
| Diarrhea, Fever, ARI, or  Enteric Virus | 232 (66.67%) | 225 (63.03%) | 226 (64.94%) | 458 (65.8%) | 451 (63.97%) | 238 (76.28%) |
| Diarrhea Fever, or ARI | 308 (41.45%) | 332 (42.29%) | 275 (35.48%) | 583 (38.41%) | 607 (38.91%) | 306 (42.21%) |
| Diarrhea | 75 (10.09%) | 62 (7.9%) | 55 (7.1%) | 130 (8.56%) | 117 (7.5%) | 84 (11.59%) |
| ARI | 145 (19.52%) | 160 (20.38%) | 128 (16.52%) | 273 (17.98%) | 288 (18.46%) | 157 (21.66%) |
| ARI with Fever  (Past 7 Days) | 73 (9.83%) | 82 (10.45%) | 75 (9.68%) | 148 (9.75%) | 157 (10.06%) | 95 (13.1%) |
| Fever (Past 7 Days) | 201 (27.05%) | 225 (28.66%) | 199 (25.68%) | 400 (26.35%) | 424 (27.18%) | 209 (28.83%) |
| Number of Enteric Viruses | 0.41 (0.64) | 0.32 (0.56) | 0.41 (0.63) | 0.41 (0.63) | 0.36 (0.59) | 0.60 (0.63) |
| Any Enteric Viruses | 116 (33.33%) | 98 (27.45%) | 116 (33.33%) | 232 (33.33%) | 214 (30.35%) | 163 (52.24%) |
| **ANTIBIOTIC OUTCOMES** | | | | | | |
| Any Antibiotics  (Past Month) | 324 (43.43%) | 350 (44.25%) | 366 (47.16%) | 690 (45.34%) | 716 (45.69%) | 363 (49.46%) |
| Any Antibiotics  (Past 3 Months) | 440 (58.74%) | 484 (61.11%) | 473 (60.64%) | 913 (59.71%) | 957 (60.88%) | 489 (66.53%) |
| Days of Antibiotic Use  (Past 3 Months) | 4.38 (5.41) | 4.32 (4.99) | 4.33 (5.12) | 4.36 (5.26) | 4.33 (5.05) | 5.17 (5.59) |
| Episodes of Antibiotic Use  (Past 3 Months) | 0.84 (0.92) | 0.86 (0.88) | 0.84 (0.87) | 0.84 (0.89) | 0.85 (0.87) | 1.00 (0.95) |
| Multiple Episodes of  Antibiotic Use  (Past 3 Months) | 133 (17.76%) | 156 (19.7%) | 138 (17.69%) | 271 (17.72%) | 294 (18.7%) | 181 (24.63%) |

Sample sizes, child demographics, household characteristics, mediator prevalence, and antibiotic use prevalence in each intervention group. For categorical variables, the number of occurrences and percentages are reported. For continuous variables, the mean and standard deviation (SD) are reported.

### Table S3: Distribution of Baseline Characteristics, Mediators, and Antibiotic Use Outcomes, By Intervention Arm and Follow-Up Period

|  | **Any WASH Intervention**  **(N = 1,080)** | | **Control**  **(N = 329)** | |
| --- | --- | --- | --- | --- |
|  | **14 Mos.**  **N=1,151** | **28 Mos.**  **N=1,170** | **14 Mos.**  **N=377** | **28 Mos**  **N=358** |
| **BASELINE COVARIATES** | | | | |
| Month of Measurement | | | | |
| *Feb-May* | 265 (23.02%) | 424 (36.24%) | 139 (36.87%) | 114 (31.84%) |
| *Jun-Sep* | 633 (55%) | 293 (25.04%) | 89 (23.61%) | 169 (47.21%) |
| *Oct-Jan* | 253 (21.98%) | 453 (38.72%) | 149 (39.52%) | 75 (20.95%) |
| Child's age | 8.56 (1.74) | 22.84 (2.15) | 9.42 (1.72) | 23.74 (1.98) |
| Child's sex | | | | |
| *Female* | 574 (49.87%) | 568 (48.55%) | 189 (50.13%) | 183 (51.12%) |
| *Male* | 577 (50.13%) | 602 (51.45%) | 188 (49.87%) | 175 (48.88%) |
| Birth Order | | | | |
| *1* | 383 (33.28%) | 403 (34.44%) | 139 (36.87%) | 133 (37.15%) |
| *2+* | 730 (63.42%) | 756 (64.62%) | 206 (54.64%) | 210 (58.66%) |
| *Missing* | 38 (3.3%) | 11 (0.94%) | 32 (8.49%) | 15 (4.19%) |
| Mother's age | 24.18 (5.33) | 24.14 (5.27) | 23.54 (4.83) | 23.37 (4.87) |
| Mother's height | 150.31 (5.44) | 150.28 (5.38) | 150.84 (5.12) | 150.83 (5.21) |
| Mother's education | | | | |
| *No education* | 168 (14.6%) | 166 (14.19%) | 43 (11.41%) | 35 (9.78%) |
| *Primary (1-5y)* | 340 (29.54%) | 352 (30.09%) | 91 (24.14%) | 88 (24.58%) |
| *Secondary (>5y)* | 643 (55.86%) | 652 (55.73%) | 243 (64.46%) | 235 (65.64%) |
| Household food insecurity | | | | |
| *Food Secure* | 807 (70.11%) | 819 (70%) | 280 (74.27%) | 263 (73.46%) |
| *Mildly Food Insecure* | 98 (8.51%) | 103 (8.8%) | 29 (7.69%) | 33 (9.22%) |
| *Moderately/Severely Food Insecure* | 246 (21.37%) | 248 (21.2%) | 68 (18.04%) | 62 (17.32%) |
| Number of individuals in household under age 18 | 1.62 (1.27) | 1.64 (1.28) | 1.55 (1.24) | 1.56 (1.27) |
| Number of individuals in compound | 11.43 (6.59) | 11.39 (6.51) | 10.05 (6.13) | 10.16 (6.01) |
| Distance to water source | 0.88 (3.94) | 0.89 (3.94) | 0.76 (1.53) | 0.75 (1.54) |
| Household Roof Material | 1136 (98.7%) | 1153 (98.55%) | 374 (99.2%) | 355 (99.16%) |
| Household Wall Material | 754 (65.51%) | 776 (66.32%) | 227 (60.21%) | 222 (62.01%) |
| Household Floor Material | 136 (11.82%) | 138 (11.79%) | 63 (16.71%) | 58 (16.2%) |
| Household Wealth Index | | | | |
| *Wealth Q1* | 273 (23.72%) | 243 (20.77%) | 53 (14.06%) | 47 (13.13%) |
| *Wealth Q2* | 285 (24.76%) | 255 (21.79%) | 72 (19.1%) | 66 (18.44%) |
| *Wealth Q3* | 247 (21.46%) | 225 (19.23%) | 99 (26.26%) | 86 (24.02%) |
| *Wealth Q4* | 274 (23.81%) | 249 (21.28%) | 105 (27.85%) | 88 (24.58%) |
| *Missing* | 72 (6.26%) | 198 (16.92%) | 48 (12.73%) | 71 (19.83%) |
| **MEDIATORS** | | | | |
| Diarrhea, Fever, ARI, or Enteric Virus | 683 (64.86%) | NA | 238 (76.28%) | NA |
| Diarrhea Fever, or ARI | 596 (51.78%) | 319 (27.69%) | 198 (52.52%) | 108 (31.03%) |
| Diarrhea | 147 (12.77%) | 45 (3.91%) | 67 (17.77%) | 17 (4.89%) |
| ARI | 347 (30.15%) | 86 (7.47%) | 119 (31.56%) | 38 (10.92%) |
| ARI with Fever | 183 (15.9%) | 47 (4.08%) | 75 (19.89%) | 20 (5.75%) |
| Fever (Past 7 Days) | 368 (31.97%) | 257 (22.31%) | 126 (33.42%) | 83 (23.85%) |
| Number of Enteric Viruses | 0.38 (0.61) | NA | 0.60 (0.63) | NA |
| Any Enteric Viruses | 330 (31.34%) | NA | 163 (52.24%) | NA |
| **ANTIBIOTIC OUTCOMES** | | | | |
| Any Antibiotics  (Past Month) | 621 (53.95%) | 419 (36.06%) | 221 (58.78%) | 142 (39.66%) |
| Any Antibiotics  (Past 3 Months) | 807 (70.11%) | 590 (50.43%) | 283 (75.07%) | 206 (57.54%) |
| Days of Antibiotic Use (Past 3 Months) | 5.39 (5.61) | 3.32 (4.47) | 6.27 (5.91) | 4.01 (4.99) |
| Episodes of Antibiotic Use  (Past 3 Months) | 1.06 (0.96) | 0.65 (0.76) | 1.23 (0.99) | 0.77 (0.84) |
| Multiple Episodes of Antibiotic Use  (Past 3 Months) | 295 (25.63%) | 132 (11.28%) | 131 (34.75%) | 50 (13.97%) |

Sample sizes, child characteristics, mediator prevalence, and antibiotic use prevalence in each intervention group, by follow-up period. For categorical variables, the number of occurrences and percentages are reported. For continuous variables, the mean and standard deviation (SD) are reported.

### Table S4: Joint Prevalence of Mediators

| **Age** | **Mediator** | **Pooled Intervention** | **Control** |
| --- | --- | --- | --- |
| Pooled Ages | Diarrhea only | 34 (1.5%) | 8 (1.1%) |
|  | ARI only | 87 (3.7%) | 13 (1.8%) |
|  | Fever only | 86 (3.7%) | 16 (2.2%) |
|  | Enteric Virus only | 138 (5.9%) | 71 (9.7%) |
|  | Diarrhea + ARI | 8 (0.3%) | 1 (0.1%) |
|  | Diarrhea + Fever | 26 (1.1%) | 4 (0.5%) |
|  | Diarrhea + ARI + Fever | 10 (0.4%) | 4 (0.5%) |
|  | ARI + Enteric Virus | 43 (1.9%) | 18 (2.4%) |
|  | Diarrhea + ARI + Fever + Enteric Virus | 14 (0.6%) | 8 (1.1%) |
| 14 months | Diarrhea only | 34 (3.0%) | 8 (2.1%) |
|  | ARI only | 87 (7.6%) | 13 (3.4%) |
|  | Fever only | 86 (7.5%) | 16 (4.2%) |
|  | Enteric Virus only | 138 (12.0%) | 71 (18.8%) |
|  | Diarrhea + ARI | 8 (0.7%) | 1 (0.3%) |
|  | Diarrhea + Fever | 26 (2.3%) | 4 (1.1%) |
|  | Diarrhea + ARI + Fever | 10 (0.9%) | 4 (1.1%) |
|  | ARI + Enteric Virus | 43 (3.7%) | 18 (4.8%) |
|  | Diarrhea + ARI + Fever + Enteric Virus | 14 (1.2%) | 8 (2.1%) |
| 28 months | Diarrhea only | 23 (2.0%) | 7 (2.0%) |
|  | ARI only | 38 (3.2%) | 16 (4.5%) |
|  | Fever only | 191 (16.3%) | 57 (15.9%) |
|  | Diarrhea + ARI | 1 (0.1%) | 2 (0.6%) |
|  | Diarrhea + Fever | 19 (1.6%) | 6 (1.7%) |
|  | Diarrhea + ARI + Fever | 2 (0.2%) | 2 (0.6%) |

Number and percentage of children with various combinations of co-occurring mediators in measurements taken at 14 and/or 28 months of age

### Table S5: Intervention-Outcome Effects

| **Intervention Group** | **Outcome** | **Prevalence/Mean in Control Arm** | **Prevalence/Mean in Intervention Arm** | **Prevalence/Mean Difference (95% CI)** | **Prevalence Ratio (95% CI)** |
| --- | --- | --- | --- | --- | --- |
| Pooled Intervention | Any antibiotic use in past month | 0.495 (0.459, 0.530) | 0.450 (0.427, 0.472) | -0.055 (-0.099, -0.012) | 0.890 (0.813, 0.974) |
|  | Any antibiotic use in past 3 months | 0.665 (0.618, 0.713) | 0.602 (0.581, 0.623) | -0.072 (-0.124, -0.017) | 0.892 (0.822, 0.967) |
|  | Days of antibiotic use in past 3 months | 5.167 (4.724, 5.610) | 4.345 (4.120, 4.571) | -0.929 (-1.405, -0.453) | ---- |
|  | Episodes of antibiotic use in past 3 months | 1.004 (0.929, 1.079) | 0.850 (0.812, 0.889) | -0.175 (-0.257, -0.092) | ---- |
|  | Multiple episodes of antibiotic use | 0.246 (0.214, 0.279) | 0.184 (0.167, 0.201) | -0.071 (-0.105, -0.038) | 0.719 (0.623, 0.829) |
| Pooled Nutrition and Nutrition + WSH | Any antibiotic use in past month | 0.495 (0.459, 0.530) | 0.453 (0.425, 0.481) | -0.050 (-0.097, -0.002) | 0.901 (0.815, 0.997) |
|  | Any antibiotic use in past 3 months | 0.665 (0.618, 0.713) | 0.597 (0.570, 0.625) | -0.074 (-0.124, -0.019) | 0.887 (0.815, 0.965) |
|  | Days of antibiotic use in past 3 months | 5.167 (4.724, 5.610) | 4.357 (4.029, 4.685) | -0.917 (-1.443, -0.390) | ---- |
|  | Episodes of antibiotic use in past 3 months | 1.004 (0.929, 1.079) | 0.843 (0.791, 0.895) | -0.181 (-0.269, -0.092) | ---- |
|  | Multiple episodes of antibiotic use | 0.246 (0.214, 0.279) | 0.177 (0.157, 0.198) | -0.077 (-0.117, -0.041) | 0.692 (0.587, 0.815) |
| Pooled WSH and Nutrition + WSH | Any antibiotic use in past month | 0.495 (0.459, 0.530) | 0.457 (0.431, 0.483) | -0.049 (-0.094, -0.005) | 0.903 (0.823, 0.991) |
|  | Any antibiotic use in past 3 months | 0.665 (0.618, 0.713) | 0.609 (0.582, 0.636) | -0.065 (-0.122, -0.009) | 0.900 (0.825, 0.981) |
|  | Days of antibiotic use in past 3 months | 5.167 (4.724, 5.610) | 4.328 (4.090, 4.567) | -0.960 (-1.454, -0.465) | ---- |
|  | Episodes of antibiotic use in past 3 months | 1.004 (0.929, 1.079) | 0.854 (0.809, 0.898) | -0.174 (-0.260, -0.088) | ---- |
|  | Multiple episodes of antibiotic use | 0.246 (0.214, 0.279) | 0.187 (0.168, 0.206) | -0.069 (-0.107, -0.035) | 0.730 (0.626, 0.851) |
| Nutrition | Any antibiotic use in past month | 0.495 (0.459, 0.530) | 0.434 (0.397, 0.471) | -0.070 (-0.123, -0.017) | 0.860 (0.765, 0.967) |
|  | Any antibiotic use in past 3 months | 0.665 (0.618, 0.713) | 0.587 (0.549, 0.626) | -0.083 (-0.143, -0.027) | 0.875 (0.795, 0.962) |
|  | Days of antibiotic use in past 3 months | 5.167 (4.724, 5.610) | 4.381 (3.935, 4.827) | -0.873 (-1.463, -0.283) | ---- |
|  | Episodes of antibiotic use in past 3 months | 1.004 (0.929, 1.079) | 0.844 (0.770, 0.917) | -0.179 (-0.280, -0.077) | ---- |
|  | Multiple episodes of antibiotic use | 0.246 (0.214, 0.279) | 0.178 (0.149, 0.206) | -0.077 (-0.117, -0.038) | 0.692 (0.574, 0.834) |
| WSH | Any antibiotic use in past month | 0.495 (0.459, 0.530) | 0.442 (0.406, 0.479) | -0.066 (-0.115, -0.020) | 0.867 (0.780, 0.964) |
|  | Any antibiotic use in past 3 months | 0.665 (0.618, 0.713) | 0.611 (0.578, 0.645) | -0.066 (-0.127, -0.003) | 0.901 (0.820, 0.990) |
|  | Days of antibiotic use in past 3 months | 5.167 (4.724, 5.610) | 4.323 (4.022, 4.624) | -0.960 (-1.481, -0.438) | ---- |
|  | Episodes of antibiotic use in past 3 months | 1.004 (0.929, 1.079) | 0.865 (0.802, 0.928) | -0.165 (-0.262, -0.067) | ---- |
|  | Multiple episodes of antibiotic use | 0.246 (0.214, 0.279) | 0.197 (0.169, 0.225) | -0.060 (-0.100, -0.021) | 0.767 (0.644, 0.913) |
| Nutrition + WSH | Any antibiotic use in past month | 0.495 (0.459, 0.530) | 0.472 (0.434, 0.509) | -0.031 (-0.086, 0.020) | 0.940 (0.841, 1.050) |
|  | Any antibiotic use in past 3 months | 0.665 (0.618, 0.713) | 0.606 (0.567, 0.646) | -0.067 (-0.129, -0.011) | 0.900 (0.817, 0.990) |
|  | Days of antibiotic use in past 3 months | 5.167 (4.724, 5.610) | 4.334 (3.948, 4.720) | -0.958 (-1.538, -0.379) | ---- |
|  | Episodes of antibiotic use in past 3 months | 1.004 (0.929, 1.079) | 0.842 (0.776, 0.909) | -0.184 (-0.281, -0.086) | ---- |
|  | Multiple episodes of antibiotic use | 0.246 (0.214, 0.279) | 0.177 (0.152, 0.202) | -0.076 (-0.118, -0.034) | 0.691 (0.569, 0.839) |

Point estimates and 95% confidence intervals (CIs) for the total effect of any WASH or nutrition intervention on antibiotic use outcomes. For categorical outcomes (any antibiotic use, multiple episodes of antibiotic use), the prevalence difference and prevalence ratio are reported. For continuous outcomes (days of antibiotic use, episodes of antibiotic use), only the mean difference is reported. Antibiotic use is reported by a caregiver under either a 1- or 3-month lookback period at 14 and 28 months.

### Table S6: Intervention-Mediator Effects

| **Intervention Group** | **Mediator** | **Prevalence/Mean in Control Arm** | **Prevalence/Mean in Intervention Arm** | **Mean Difference (95% CI)** | **Prevalence Ratio (95% CI)** |
| --- | --- | --- | --- | --- | --- |
| Pooled Intervention | Diarrhea, Fever, ARI, or Enteric Virus | 0.763 (0.714, 0.812) | 0.649 (0.617, 0.680) | ---- | 0.844 (0.773, 0.921) |
|  | Diarrhea, Fever, or ARI | 0.422 (0.390, 0.454) | 0.397 (0.378, 0.417) | ---- | 0.913 (0.833, 1.001) |
|  | Diarrhea | 0.116 (0.092, 0.140) | 0.083 (0.069, 0.097) | ---- | 0.671 (0.508, 0.886) |
|  | ARI | 0.217 (0.183, 0.250) | 0.188 (0.172, 0.204) | ---- | 0.835 (0.702, 0.994) |
|  | ARI with Fever | 0.131 (0.107, 0.155) | 0.100 (0.088, 0.112) | ---- | 0.739 (0.604, 0.906) |
|  | Difficulty Breathing | 0.040 (0.026, 0.054) | 0.029 (0.022, 0.037) | ---- | 0.706 (0.449, 1.110) |
|  | Fever (Past 7 Days) | 0.288 (0.254, 0.323) | 0.271 (0.253, 0.290) | ---- | 0.925 (0.819, 1.044) |
|  | Fever (Past 14 Days) | 0.443 (0.391, 0.495) | 0.421 (0.388, 0.453) | ---- | 0.936 (0.810, 1.081) |
|  | Any Enteric Virus | 0.522 (0.451, 0.594) | 0.313 (0.277, 0.350) | ---- | 0.579 (0.492, 0.682) |
|  | Any Enteric Virus with Diarrhea | 0.063 (0.041, 0.084) | 0.027 (0.021, 0.033) | ---- | 0.351 (0.241, 0.510) |
|  | Number of Enteric Viruses | 0.599 (0.501, 0.698) | 0.377 (0.327, 0.427) | -0.238 (-0.341, -0.135) | ---- |
|  | Adenovirus 40/41 | 0.104 (0.048, 0.160) | 0.070 (0.047, 0.094) | ---- | 0.602 (0.352, 1.027) |
|  | Norovirus GII | 0.191 (0.146, 0.237) | 0.133 (0.107, 0.160) | ---- | 0.669 (0.481, 0.930) |
|  | Sapovirus | 0.195 (0.142, 0.248) | 0.101 (0.080, 0.122) | ---- | 0.507 (0.356, 0.721) |
| Pooled Nutrition and Nutrition + WSH | Diarrhea, Fever, ARI, or Enteric Virus | 0.763 (0.714, 0.812) | 0.658 (0.620, 0.696) | ---- | 0.854 (0.781, 0.934) |
|  | Diarrhea, Fever, or ARI | 0.422 (0.390, 0.454) | 0.384 (0.359, 0.409) | ---- | 0.883 (0.799, 0.976) |
|  | Diarrhea | 0.116 (0.092, 0.140) | 0.086 (0.068, 0.103) | ---- | 0.689 (0.514, 0.924) |
|  | ARI | 0.217 (0.183, 0.250) | 0.180 (0.160, 0.199) | ---- | 0.803 (0.656, 0.982) |
|  | ARI with Fever | 0.131 (0.107, 0.155) | 0.097 (0.083, 0.112) | ---- | 0.726 (0.573, 0.921) |
|  | Difficulty Breathing | 0.040 (0.026, 0.054) | 0.030 (0.020, 0.039) | ---- | 0.719 (0.452, 1.143) |
|  | Fever (Past 7 Days) | 0.288 (0.254, 0.323) | 0.264 (0.241, 0.286) | ---- | 0.899 (0.790, 1.024) |
|  | Fever (Past 14 Days) | 0.443 (0.391, 0.495) | 0.409 (0.371, 0.447) | ---- | 0.900 (0.778, 1.041) |
|  | Any Enteric Virus | 0.522 (0.451, 0.594) | 0.333 (0.288, 0.379) | ---- | 0.623 (0.524, 0.739) |
|  | Any Enteric Virus with Diarrhea | 0.063 (0.041, 0.084) | 0.028 (0.021, 0.035) | ---- | 0.368 (0.247, 0.549) |
|  | Number of Enteric Viruses | 0.599 (0.501, 0.698) | 0.408 (0.345, 0.471) | -0.202 (-0.307, -0.098) | ---- |
|  | Adenovirus 40/41 | 0.104 (0.048, 0.160) | 0.077 (0.048, 0.106) | ---- | 0.667 (0.391, 1.140) |
|  | Norovirus GII | 0.191 (0.146, 0.237) | 0.156 (0.121, 0.192) | ---- | 0.799 (0.570, 1.119) |
|  | Sapovirus | 0.195 (0.142, 0.248) | 0.109 (0.085, 0.134) | ---- | 0.554 (0.378, 0.814) |
| Pooled WSH and Nutrition + WSH | Diarrhea, Fever, ARI, or Enteric Virus | 0.763 (0.714, 0.812) | 0.640 (0.603, 0.677) | ---- | 0.829 (0.751, 0.916) |
|  | Diarrhea, Fever, or ARI | 0.422 (0.390, 0.454) | 0.389 (0.366, 0.412) | ---- | 0.894 (0.812, 0.985) |
|  | Diarrhea | 0.116 (0.092, 0.140) | 0.075 (0.059, 0.091) | ---- | 0.605 (0.441, 0.829) |
|  | ARI | 0.217 (0.183, 0.250) | 0.185 (0.164, 0.205) | ---- | 0.818 (0.690, 0.968) |
|  | ARI with Fever | 0.131 (0.107, 0.155) | 0.101 (0.086, 0.116) | ---- | 0.744 (0.605, 0.915) |
|  | Difficulty Breathing | 0.040 (0.026, 0.054) | 0.027 (0.018, 0.036) | ---- | 0.649 (0.393, 1.070) |
|  | Fever (Past 7 Days) | 0.288 (0.254, 0.323) | 0.272 (0.248, 0.296) | ---- | 0.927 (0.814, 1.055) |
|  | Fever (Past 14 Days) | 0.443 (0.391, 0.495) | 0.425 (0.384, 0.466) | ---- | 0.964 (0.827, 1.124) |
|  | Any Enteric Virus | 0.522 (0.451, 0.594) | 0.304 (0.264, 0.344) | ---- | 0.559 (0.465, 0.673) |
|  | Any Enteric Virus with Diarrhea | 0.063 (0.041, 0.084) | 0.024 (0.016, 0.031) | ---- | 0.302 (0.193, 0.473) |
|  | Number of Enteric Viruses | 0.599 (0.501, 0.698) | 0.360 (0.305, 0.415) | -0.262 (-0.375, -0.148) | ---- |
|  | Adenovirus 40/41 | 0.104 (0.048, 0.160) | 0.064 (0.041, 0.087) | ---- | 0.513 (0.279, 0.945) |
|  | Norovirus GII | 0.191 (0.146, 0.237) | 0.121 (0.093, 0.149) | ---- | 0.610 (0.423, 0.879) |
|  | Sapovirus | 0.195 (0.142, 0.248) | 0.102 (0.075, 0.129) | ---- | 0.509 (0.353, 0.734) |
| Nutrition | Diarrhea, Fever, ARI, or Enteric Virus | 0.763 (0.714, 0.812) | 0.667 (0.614, 0.720) | ---- | 0.865 (0.783, 0.955) |
|  | Diarrhea, Fever, or ARI | 0.422 (0.390, 0.454) | 0.415 (0.381, 0.448) | ---- | 0.955 (0.853, 1.070) |
|  | Diarrhea | 0.116 (0.092, 0.140) | 0.101 (0.074, 0.128) | ---- | 0.807 (0.581, 1.121) |
|  | ARI | 0.217 (0.183, 0.250) | 0.195 (0.170, 0.220) | ---- | 0.877 (0.696, 1.105) |
|  | ARI with Fever | 0.131 (0.107, 0.155) | 0.098 (0.079, 0.117) | ---- | 0.733 (0.555, 0.966) |
|  | Difficulty Breathing | 0.040 (0.026, 0.054) | 0.034 (0.021, 0.046) | ---- | 0.824 (0.481, 1.411) |
|  | Fever (Past 7 Days) | 0.288 (0.254, 0.323) | 0.271 (0.238, 0.303) | ---- | 0.922 (0.785, 1.082) |
|  | Fever (Past 14 Days) | 0.443 (0.391, 0.495) | 0.411 (0.363, 0.460) | ---- | 0.899 (0.763, 1.059) |
|  | Any Enteric Virus | 0.522 (0.451, 0.594) | 0.333 (0.264, 0.403) | ---- | 0.615 (0.490, 0.772) |
|  | Any Enteric Virus with Diarrhea | 0.063 (0.041, 0.084) | 0.034 (0.022, 0.046) | ---- | 0.450 (0.288, 0.704) |
|  | Number of Enteric Viruses | 0.599 (0.501, 0.698) | 0.411 (0.319, 0.503) | -0.204 (-0.325, -0.082) | ---- |
|  | Adenovirus 40/41 | 0.104 (0.048, 0.160) | 0.083 (0.043, 0.123) | ---- | 0.739 (0.403, 1.355) |
|  | Norovirus GII | 0.191 (0.146, 0.237) | 0.159 (0.110, 0.208) | ---- | 0.806 (0.543, 1.196) |
|  | Sapovirus | 0.195 (0.142, 0.248) | 0.100 (0.062, 0.137) | ---- | 0.483 (0.284, 0.821) |
| WSH | Diarrhea, Fever, ARI, or Enteric Virus | 0.763 (0.714, 0.812) | 0.630 (0.582, 0.679) | ---- | 0.812 (0.725, 0.908) |
|  | Diarrhea, Fever, or ARI | 0.422 (0.390, 0.454) | 0.423 (0.395, 0.451) | ---- | 0.972 (0.876, 1.078) |
|  | Diarrhea | 0.116 (0.092, 0.140) | 0.079 (0.060, 0.098) | ---- | 0.633 (0.450, 0.889) |
|  | ARI | 0.217 (0.183, 0.250) | 0.204 (0.179, 0.229) | ---- | 0.903 (0.766, 1.066) |
|  | ARI with Fever | 0.131 (0.107, 0.155) | 0.104 (0.083, 0.126) | ---- | 0.768 (0.609, 0.968) |
|  | Difficulty Breathing | 0.040 (0.026, 0.054) | 0.028 (0.016, 0.040) | ---- | 0.679 (0.372, 1.238) |
|  | Fever (Past 7 Days) | 0.288 (0.254, 0.323) | 0.287 (0.256, 0.317) | ---- | 0.975 (0.842, 1.128) |
|  | Fever (Past 14 Days) | 0.443 (0.391, 0.495) | 0.443 (0.390, 0.496) | ---- | 1.010 (0.851, 1.200) |
|  | Any Enteric Virus | 0.522 (0.451, 0.594) | 0.275 (0.225, 0.324) | ---- | 0.491 (0.389, 0.619) |
|  | Any Enteric Virus with Diarrhea | 0.063 (0.041, 0.084) | 0.026 (0.016, 0.037) | ---- | 0.317 (0.190, 0.530) |
|  | Number of Enteric Viruses | 0.599 (0.501, 0.698) | 0.317 (0.251, 0.382) | -0.320 (-0.444, -0.197) | ---- |
|  | Adenovirus 40/41 | 0.104 (0.048, 0.160) | 0.058 (0.031, 0.085) | ---- | 0.452 (0.220, 0.928) |
|  | Norovirus GII | 0.191 (0.146, 0.237) | 0.090 (0.061, 0.119) | ---- | 0.437 (0.288, 0.664) |
|  | Sapovirus | 0.195 (0.142, 0.248) | 0.085 (0.052, 0.119) | ---- | 0.403 (0.252, 0.644) |
| Nutrition + WSH | Diarrhea, Fever, ARI, or Enteric Virus | 0.763 (0.714, 0.812) | 0.649 (0.596, 0.703) | ---- | 0.837 (0.748, 0.937) |
|  | Diarrhea, Fever, or ARI | 0.422 (0.390, 0.454) | 0.355 (0.318, 0.392) | ---- | 0.817 (0.719, 0.928) |
|  | Diarrhea | 0.116 (0.092, 0.140) | 0.071 (0.049, 0.093) | ---- | 0.575 (0.392, 0.844) |
|  | ARI | 0.217 (0.183, 0.250) | 0.165 (0.135, 0.195) | ---- | 0.737 (0.585, 0.928) |
|  | ARI with Fever | 0.131 (0.107, 0.155) | 0.097 (0.077, 0.117) | ---- | 0.724 (0.553, 0.948) |
|  | Difficulty Breathing | 0.040 (0.026, 0.054) | 0.026 (0.012, 0.039) | ---- | 0.621 (0.340, 1.135) |
|  | Fever (Past 7 Days) | 0.288 (0.254, 0.323) | 0.257 (0.224, 0.289) | ---- | 0.880 (0.754, 1.026) |
|  | Fever (Past 14 Days) | 0.443 (0.391, 0.495) | 0.406 (0.351, 0.462) | ---- | 0.916 (0.773, 1.085) |
|  | Any Enteric Virus | 0.522 (0.451, 0.594) | 0.333 (0.273, 0.393) | ---- | 0.624 (0.504, 0.773) |
|  | Any Enteric Virus with Diarrhea | 0.063 (0.041, 0.084) | 0.021 (0.011, 0.031) | ---- | 0.287 (0.162, 0.507) |
|  | Number of Enteric Viruses | 0.599 (0.501, 0.698) | 0.405 (0.323, 0.487) | -0.212 (-0.335, -0.089) | ---- |
|  | Adenovirus 40/41 | 0.104 (0.048, 0.160) | 0.071 (0.038, 0.103) | ---- | 0.571 (0.299, 1.089) |
|  | Norovirus GII | 0.191 (0.146, 0.237) | 0.153 (0.109, 0.197) | ---- | 0.795 (0.533, 1.186) |
|  | Sapovirus | 0.195 (0.142, 0.248) | 0.119 (0.080, 0.157) | ---- | 0.606 (0.396, 0.927) |

Point estimates and 95% confidence intervals for the effects of WASH and/or Nutrition interventions on potential mediators. For categorical mediators, the prevalence ratio is reported. For continuous mediators, the mean difference is reported. Diarrhea, ARI, ARI with Fever, and Fever are all reported by a caregiver under a 7-day lookback period at 14 and 28 months. Any Enteric Virus is the presence of adenovirus 40/41, norovirus GI, norovirus GII, sapovirus, rotavirus, or astrovirus in stool collected at 14 months, and Any Enteric Virus with Diarrhea is the presence of any enteric virus with caregiver reported diarrhea in the prior 7 days at 14 months.

### Table S7: Mediator-Outcome Effects

| **Mediator** | **Outcome** | **Prevalence/Mean in Absence of Mediator** | **Prevalence/Mean in Presence of Mediator** | **Mean Difference (95% CI)** | **Prevalence Ratio (95% CI)** |
| --- | --- | --- | --- | --- | --- |
| Diarrhea, Fever, ARI, or Enteric Virus | Any antibiotic use in past month | 0.426 (0.387, 0.465) | 0.618 (0.588, 0.649) | ---- | 1.435 (1.299, 1.585) |
| Diarrhea, Fever, or ARI | Any antibiotic use in past month | 0.395 (0.374, 0.416) | 0.559 (0.525, 0.593) | ---- | 1.311 (1.220, 1.409) |
| Diarrhea | Any antibiotic use in past month | 0.444 (0.424, 0.465) | 0.630 (0.569, 0.692) | ---- | 1.291 (1.170, 1.425) |
| ARI | Any antibiotic use in past month | 0.423 (0.402, 0.444) | 0.620 (0.575, 0.664) | ---- | 1.326 (1.222, 1.440) |
| ARI with Fever | Any antibiotic use in past month | 0.435 (0.414, 0.456) | 0.679 (0.623, 0.735) | ---- | 1.407 (1.283, 1.542) |
| Difficulty Breathing | Any antibiotic use in past month | 0.454 (0.433, 0.474) | 0.695 (0.595, 0.794) | ---- | 1.470 (1.274, 1.696) |
| Fever (Past 7 Days) | Any antibiotic use in past month | 0.422 (0.401, 0.443) | 0.564 (0.527, 0.602) | ---- | 1.268 (1.176, 1.368) |
| Fever (Past 14 Days) | Any antibiotic use in past month | 0.458 (0.427, 0.490) | 0.677 (0.635, 0.719) | ---- | 1.475 (1.337, 1.626) |
| Any Enteric Virus | Any antibiotic use in past month | 0.521 (0.485, 0.556) | 0.618 (0.578, 0.658) | ---- | 1.168 (1.058, 1.288) |
|  | Any antibiotic use in past 3 months | 0.698 (0.664, 0.733) | 0.753 (0.714, 0.791) | ---- | 1.080 (1.003, 1.162) |
|  | Days of antibiotic use in past 3 months | 5.536 (5.180, 5.893) | 5.804 (5.276, 6.333) | 0.231 (-0.380, 0.843) | ---- |
|  | Episodes of antibiotic use in past 3 months | 1.076 (1.010, 1.142) | 1.170 (1.083, 1.257) | 0.077 (-0.033, 0.187) | ---- |
|  | Multiple episodes of antibiotic use | 0.269 (0.240, 0.299) | 0.308 (0.264, 0.353) | ---- | 1.103 (0.925, 1.316) |
| Any Enteric Virus with Diarrhea | Any antibiotic use in past month | 0.452 (0.431, 0.472) | 0.752 (0.667, 0.838) | ---- | 1.421 (1.249, 1.616) |
|  | Any antibiotic use in past 3 months | 0.610 (0.589, 0.632) | 0.829 (0.757, 0.900) | ---- | 1.179 (1.075, 1.292) |
|  | Days of antibiotic use in past 3 months | 4.484 (4.258, 4.709) | 6.625 (5.391, 7.859) | 1.158 (-0.061, 2.377) | ---- |
|  | Episodes of antibiotic use in past 3 months | 0.874 (0.835, 0.914) | 1.343 (1.164, 1.522) | 0.265 (0.076, 0.453) | ---- |
|  | Multiple episodes of antibiotic use | 0.195 (0.177, 0.213) | 0.362 (0.273, 0.451) | ---- | 1.325 (1.006, 1.745) |
| Number of Enteric Viruses | Any antibiotic use in past month | 0.521 (0.485, 0.556) | 0.618 (0.578, 0.658) | ---- | 1.085 (1.010, 1.166) |
|  | Any antibiotic use in past 3 months | 0.698 (0.664, 0.733) | 0.753 (0.714, 0.791) | ---- | 1.031 (0.976, 1.090) |
|  | Days of antibiotic use in past 3 months | 5.536 (5.180, 5.893) | 5.804 (5.276, 6.333) | 0.018 (-0.462, 0.498) | ---- |
|  | Episodes of antibiotic use in past 3 months | 1.076 (1.010, 1.142) | 1.170 (1.083, 1.257) | 0.020 (-0.061, 0.102) | ---- |
|  | Multiple episodes of antibiotic use | 0.269 (0.240, 0.299) | 0.308 (0.264, 0.353) | ---- | 1.030 (0.902, 1.176) |
| Adenovirus 40/41 | Any antibiotic use in past month | 0.557 (0.528, 0.585) | 0.569 (0.476, 0.661) | ---- | 1.026 (0.864, 1.217) |
|  | Any antibiotic use in past 3 months | 0.720 (0.693, 0.747) | 0.725 (0.660, 0.789) | ---- | 1.006 (0.907, 1.116) |
|  | Days of antibiotic use in past 3 months | 5.684 (5.375, 5.993) | 5.444 (4.518, 6.370) | -0.129 (-1.118, 0.861) | ---- |
|  | Episodes of antibiotic use in past 3 months | 1.107 (1.055, 1.159) | 1.183 (0.991, 1.376) | 0.078 (-0.122, 0.278) | ---- |
|  | Multiple episodes of antibiotic use | 0.282 (0.257, 0.307) | 0.312 (0.202, 0.422) | ---- | 1.119 (0.793, 1.578) |
| Norovirus GII | Any antibiotic use in past month | 0.551 (0.522, 0.580) | 0.609 (0.542, 0.676) | ---- | 1.089 (0.956, 1.240) |
|  | Any antibiotic use in past 3 months | 0.719 (0.693, 0.746) | 0.734 (0.680, 0.789) | ---- | 1.018 (0.932, 1.112) |
|  | Days of antibiotic use in past 3 months | 5.666 (5.370, 5.963) | 5.728 (4.937, 6.519) | 0.012 (-0.810, 0.833) | ---- |
|  | Episodes of antibiotic use in past 3 months | 1.114 (1.061, 1.167) | 1.121 (1.000, 1.241) | -0.018 (-0.153, 0.117) | ---- |
|  | Multiple episodes of antibiotic use | 0.282 (0.257, 0.308) | 0.295 (0.229, 0.360) | ---- | 1.003 (0.794, 1.268) |
| Sapovirus | Any antibiotic use in past month | 0.550 (0.522, 0.579) | 0.613 (0.551, 0.675) | ---- | 1.104 (0.980, 1.245) |
|  | Any antibiotic use in past 3 months | 0.715 (0.690, 0.741) | 0.751 (0.688, 0.815) | ---- | 1.058 (0.967, 1.157) |
|  | Days of antibiotic use in past 3 months | 5.653 (5.332, 5.975) | 5.663 (4.763, 6.562) | 0.082 (-0.860, 1.023) | ---- |
|  | Episodes of antibiotic use in past 3 months | 1.110 (1.053, 1.166) | 1.142 (1.010, 1.274) | 0.030 (-0.116, 0.176) | ---- |
|  | Multiple episodes of antibiotic use | 0.281 (0.254, 0.308) | 0.302 (0.232, 0.371) | ---- | 1.062 (0.829, 1.362) |

Point estimates and 95% confidence intervals for the effects of mediators on antibiotic use. For categorical outcomes (any antibiotic use, multiple episodes of antibiotic use), the prevalence ratio is reported. For continuous outcomes (days of antibiotic use, episodes of antibiotic use), the mean difference is reported. Diarrhea, ARI, ARI with Fever, and Fever are all reported by a caregiver under a 7-day lookback period at 14 and 28 months. Any Enteric Virus is the presence of adenovirus 40/41, norovirus GI, norovirus GII, sapovirus, rotavirus, or astrovirus in stool collected at 14 months, and Any Enteric Virus with Diarrhea is the presence of any enteric virus with caregiver reported diarrhea in the prior 7 days at 14 months. Antibiotic use is reported by a caregiver under either a 1- or 3-month lookback period at 14 and 28 months.

### Table S8: Mediated Effects

| **Intervention Group** | **Mediator** | **Outcome** | **Total Natural Indirect Effect (Prevalence Difference)** | **Total Natural Indirect Effect (Prevalence Ratio)** |
| --- | --- | --- | --- | --- |
| Pooled Intervention | Diarrhea, Fever, ARI, or Enteric Virus | Any antibiotic use in past month | -0.025 (-0.053, -0.002) | 1.058 (1.012, 1.120) |
|  | Diarrhea, Fever, or ARI | Any antibiotic use in past month | -0.005 (-0.014, 0.003) | 1.021 (1.002, 1.045) |
|  | Diarrhea | Any antibiotic use in past month | -0.006 (-0.013, -0.001) | 0.997 (0.988, 1.005) |
|  | ARI | Any antibiotic use in past month | -0.006 (-0.014, 0.002) | 1.006 (0.992, 1.022) |
|  | ARI with Fever | Any antibiotic use in past month | -0.007 (-0.015, -0.001) | 1.002 (0.992, 1.014) |
|  | Difficulty Breathing | Any antibiotic use in past month | -0.003 (-0.009, 0.002) | 1.001 (0.994, 1.009) |
|  | Fever (Past 7 Days) | Any antibiotic use in past month | -0.003 (-0.008, 0.003) | 1.011 (0.999, 1.025) |
|  | Fever (Past 14 Days) | Any antibiotic use in past month | -0.007 (-0.030, 0.014) | 1.058 (1.017, 1.106) |
|  | Any Enteric Virus | Any antibiotic use in past month | -0.018 (-0.035, -0.005) | 0.982 (0.965, 0.996) |
|  |  | Any antibiotic use in past 3 months | -0.012 (-0.028, 0.001) | 0.987 (0.970, 1.000) |
|  | Any Enteric Virus with Diarrhea | Any antibiotic use in past month | -0.011 (-0.021, -0.004) | 0.989 (0.980, 0.997) |
|  |  | Any antibiotic use in past 3 months | -0.007 (-0.015, -0.002) | 0.992 (0.985, 0.997) |
|  |  | Episodes of antibiotic use in past 3 months | -0.016 (-0.033, -0.003) | ---- |
|  |  | Multiple episodes of antibiotic use | -0.004 (-0.011, 0.001) | 0.992 (0.981, 1.003) |
|  | Number of Enteric Viruses | Any antibiotic use in past month | -0.009 (-0.020, 0.000) | 0.985 (0.966, 0.999) |
| Pooled Nutrition and Nutrition + WSH | Diarrhea, Fever, ARI, or Enteric Virus | Any antibiotic use in past month | -0.013 (-0.035, 0.002) | 1.021 (0.991, 1.067) |
|  | Diarrhea, Fever, or ARI | Any antibiotic use in past month | -0.005 (-0.013, 0.001) | 1.009 (0.996, 1.027) |
|  | Diarrhea | Any antibiotic use in past month | -0.005 (-0.012, 0.000) | 0.997 (0.988, 1.007) |
|  | ARI | Any antibiotic use in past month | -0.004 (-0.011, 0.001) | 1.003 (0.994, 1.016) |
|  | ARI with Fever | Any antibiotic use in past month | -0.005 (-0.013, 0.001) | 1.001 (0.991, 1.013) |
|  | Difficulty Breathing | Any antibiotic use in past month | -0.002 (-0.008, 0.002) | 1.001 (0.993, 1.011) |
|  | Fever (Past 7 Days) | Any antibiotic use in past month | -0.003 (-0.008, 0.002) | 1.005 (0.996, 1.019) |
|  | Fever (Past 14 Days) | Any antibiotic use in past month | -0.009 (-0.029, 0.010) | 1.035 (1.001, 1.082) |
|  | Any Enteric Virus | Any antibiotic use in past month | -0.013 (-0.030, -0.002) | 0.986 (0.968, 0.999) |
|  |  | Any antibiotic use in past 3 months | -0.007 (-0.022, 0.004) | 0.992 (0.978, 1.005) |
|  | Any Enteric Virus with Diarrhea | Any antibiotic use in past month | -0.009 (-0.018, -0.002) | 0.992 (0.982, 1.000) |
|  |  | Any antibiotic use in past 3 months | -0.007 (-0.016, -0.002) | 0.992 (0.983, 0.998) |
|  |  | Episodes of antibiotic use in past 3 months | -0.010 (-0.027, 0.003) | ---- |
|  |  | Multiple episodes of antibiotic use | -0.001 (-0.008, 0.004) | 0.998 (0.983, 1.020) |
|  | Number of Enteric Viruses | Any antibiotic use in past month | -0.006 (-0.016, 0.002) | 0.990 (0.972, 1.004) |
| Pooled WSH and Nutrition + WSH | Diarrhea, Fever, ARI, or Enteric Virus | Any antibiotic use in past month | -0.041 (-0.085, -0.006) | 1.074 (1.006, 1.169) |
|  | Diarrhea, Fever, or ARI | Any antibiotic use in past month | -0.010 (-0.023, 0.001) | 1.022 (1.000, 1.049) |
|  | Diarrhea | Any antibiotic use in past month | -0.009 (-0.018, -0.002) | 0.993 (0.981, 1.003) |
|  | ARI | Any antibiotic use in past month | -0.008 (-0.019, 0.000) | 1.004 (0.990, 1.020) |
|  | ARI with Fever | Any antibiotic use in past month | -0.008 (-0.017, -0.001) | 1.000 (0.988, 1.014) |
|  | Difficulty Breathing | Any antibiotic use in past month | -0.004 (-0.012, 0.000) | 0.998 (0.989, 1.007) |
|  | Fever (Past 7 Days) | Any antibiotic use in past month | -0.005 (-0.014, 0.002) | 1.012 (0.997, 1.032) |
|  | Fever (Past 14 Days) | Any antibiotic use in past month | -0.004 (-0.030, 0.019) | 1.065 (1.017, 1.134) |
|  | Any Enteric Virus | Any antibiotic use in past month | -0.023 (-0.045, -0.007) | 0.975 (0.954, 0.991) |
|  |  | Any antibiotic use in past 3 months | -0.013 (-0.032, 0.002) | 0.986 (0.968, 1.002) |
|  | Any Enteric Virus with Diarrhea | Any antibiotic use in past month | -0.013 (-0.027, -0.005) | 0.985 (0.974, 0.995) |
|  |  | Any antibiotic use in past 3 months | -0.008 (-0.016, -0.002) | 0.992 (0.983, 0.998) |
|  |  | Episodes of antibiotic use in past 3 months | -0.020 (-0.044, -0.004) | ---- |
|  |  | Multiple episodes of antibiotic use | -0.006 (-0.015, 0.000) | 0.989 (0.974, 1.002) |
|  | Number of Enteric Viruses | Any antibiotic use in past month | -0.013 (-0.025, -0.003) | 0.977 (0.954, 0.994) |
| Nutrition | Diarrhea, Fever, ARI, or Enteric Virus | Any antibiotic use in past month | -0.011 (-0.032, 0.003) | 1.018 (0.988, 1.071) |
|  | Diarrhea, Fever, or ARI | Any antibiotic use in past month | -0.002 (-0.010, 0.006) | 1.014 (0.998, 1.040) |
|  | Diarrhea | Any antibiotic use in past month | -0.002 (-0.009, 0.003) | 1.001 (0.992, 1.014) |
|  | ARI | Any antibiotic use in past month | -0.002 (-0.009, 0.004) | 1.005 (0.994, 1.022) |
|  | ARI with Fever | Any antibiotic use in past month | -0.004 (-0.012, 0.002) | 1.001 (0.990, 1.015) |
|  | Difficulty Breathing | Any antibiotic use in past month | -0.001 (-0.006, 0.003) | 1.002 (0.995, 1.012) |
|  | Fever (Past 7 Days) | Any antibiotic use in past month | -0.002 (-0.009, 0.005) | 1.007 (0.995, 1.026) |
|  | Fever (Past 14 Days) | Any antibiotic use in past month | -0.010 (-0.034, 0.011) | 1.035 (0.998, 1.084) |
|  | Any Enteric Virus | Any antibiotic use in past month | -0.014 (-0.036, 0.000) | 0.985 (0.961, 1.004) |
|  |  | Any antibiotic use in past 3 months | -0.005 (-0.021, 0.009) | 0.995 (0.977, 1.013) |
|  | Any Enteric Virus with Diarrhea | Any antibiotic use in past month | -0.008 (-0.018, -0.001) | 0.993 (0.983, 1.001) |
|  |  | Any antibiotic use in past 3 months | -0.007 (-0.015, -0.001) | 0.993 (0.984, 0.999) |
|  |  | Episodes of antibiotic use in past 3 months | -0.008 (-0.022, 0.002) | ---- |
|  |  | Multiple episodes of antibiotic use | -0.002 (-0.008, 0.002) | 0.996 (0.983, 1.012) |
|  | Number of Enteric Viruses | Any antibiotic use in past month | -0.005 (-0.019, 0.006) | 0.991 (0.966, 1.010) |
| WSH | Diarrhea, Fever, ARI, or Enteric Virus | Any antibiotic use in past month | -0.055 (-0.131, -0.009) | 1.105 (1.006, 1.256) |
|  | Diarrhea, Fever, or ARI | Any antibiotic use in past month | -0.004 (-0.018, 0.008) | 1.042 (1.011, 1.085) |
|  | Diarrhea | Any antibiotic use in past month | -0.007 (-0.017, -0.001) | 0.996 (0.983, 1.009) |
|  | ARI | Any antibiotic use in past month | -0.005 (-0.016, 0.003) | 1.007 (0.992, 1.027) |
|  | ARI with Fever | Any antibiotic use in past month | -0.007 (-0.016, 0.001) | 1.000 (0.987, 1.018) |
|  | Difficulty Breathing | Any antibiotic use in past month | -0.002 (-0.008, 0.001) | 0.998 (0.991, 1.005) |
|  | Fever (Past 7 Days) | Any antibiotic use in past month | -0.003 (-0.014, 0.007) | 1.022 (1.000, 1.049) |
|  | Fever (Past 14 Days) | Any antibiotic use in past month | 0.001 (-0.031, 0.032) | 1.092 (1.027, 1.177) |
|  | Any Enteric Virus | Any antibiotic use in past month | -0.027 (-0.059, -0.005) | 0.968 (0.940, 0.992) |
|  |  | Any antibiotic use in past 3 months | -0.017 (-0.042, 0.003) | 0.982 (0.960, 1.003) |
|  | Any Enteric Virus with Diarrhea | Any antibiotic use in past month | -0.011 (-0.025, -0.003) | 0.987 (0.975, 0.996) |
|  |  | Any antibiotic use in past 3 months | -0.008 (-0.018, -0.001) | 0.992 (0.983, 0.998) |
|  |  | Episodes of antibiotic use in past 3 months | -0.024 (-0.049, -0.006) | ---- |
|  |  | Multiple episodes of antibiotic use | -0.008 (-0.019, -0.001) | 0.987 (0.971, 1.000) |
|  | Number of Enteric Viruses | Any antibiotic use in past month | -0.015 (-0.035, 0.000) | 0.975 (0.944, 1.004) |
| Nutrition + WSH | Diarrhea, Fever, ARI, or Enteric Virus | Any antibiotic use in past month | -0.017 (-0.050, 0.004) | 1.027 (0.988, 1.094) |
|  | Diarrhea, Fever, or ARI | Any antibiotic use in past month | -0.010 (-0.022, 0.000) | 1.005 (0.988, 1.029) |
|  | Diarrhea | Any antibiotic use in past month | -0.009 (-0.020, -0.001) | 0.993 (0.982, 1.004) |
|  | ARI | Any antibiotic use in past month | -0.004 (-0.012, 0.001) | 0.999 (0.989, 1.011) |
|  | ARI with Fever | Any antibiotic use in past month | -0.004 (-0.013, 0.002) | 1.000 (0.990, 1.013) |
|  | Difficulty Breathing | Any antibiotic use in past month | -0.003 (-0.011, 0.002) | 0.999 (0.991, 1.010) |
|  | Fever (Past 7 Days) | Any antibiotic use in past month | -0.004 (-0.014, 0.003) | 1.007 (0.994, 1.026) |
|  | Fever (Past 14 Days) | Any antibiotic use in past month | -0.008 (-0.033, 0.015) | 1.037 (0.998, 1.100) |
|  | Any Enteric Virus | Any antibiotic use in past month | -0.013 (-0.037, 0.000) | 0.988 (0.966, 1.005) |
|  |  | Any antibiotic use in past 3 months | -0.010 (-0.030, 0.004) | 0.990 (0.973, 1.008) |
|  | Any Enteric Virus with Diarrhea | Any antibiotic use in past month | -0.008 (-0.019, -0.002) | 0.991 (0.982, 0.999) |
|  |  | Any antibiotic use in past 3 months | -0.008 (-0.018, -0.001) | 0.991 (0.982, 0.999) |
|  |  | Episodes of antibiotic use in past 3 months | -0.017 (-0.043, 0.004) | ---- |
|  |  | Multiple episodes of antibiotic use | -0.002 (-0.011, 0.005) | 0.997 (0.978, 1.024) |
|  | Number of Enteric Viruses | Any antibiotic use in past month | -0.007 (-0.022, 0.001) | 0.987 (0.965, 1.002) |

Point estimates and 95% confidence intervals for the total natural indirect effects of WASH and/or Nutrition intervention on antibiotic use. For categorical outcomes (any antibiotic use, multiple episodes of antibiotic use), the prevalence ratio and prevalence difference are reported. For continuous outcomes (days of antibiotic use, episodes of antibiotic use), the mean difference is reported. Diarrhea, ARI, ARI with Fever, and Fever are all reported by a caregiver under a 7-day lookback period at 14 and 28 months. Any Enteric Virus is the presence of adenovirus 40/41, norovirus GI, norovirus GII, sapovirus, rotavirus, or astrovirus in stool collected at 14 months, and Any Enteric Virus with Diarrhea is the presence of any enteric virus with caregiver reported diarrhea in the prior 7 days at 14 months. Antibiotic use is reported by a caregiver under either a 1- or 3-month lookback period at 14 and 28 months.

### Table S9: Mediated Effects, with Intervention-Mediator Interactions

| **Intervention Group** | **Mediator** | **Outcome** | **Pure Natural Indirect Effect** | **Total Natural Indirect Effect** |
| --- | --- | --- | --- | --- |
| Pooled Intervention | Diarrhea, Fever, ARI, or Enteric Virus | Any antibiotic use in past month | -0.016 (-0.044, 0.001) | -0.028 (-0.060, -0.001) |
|  | Diarrhea, Fever, or ARI | Any antibiotic use in past month | -0.006 (-0.017, 0.002) | -0.006 (-0.015, 0.002) |
|  | Diarrhea | Any antibiotic use in past month | -0.006 (-0.014, -0.001) | -0.006 (-0.013, -0.001) |
|  | ARI | Any antibiotic use in past month | -0.003 (-0.010, 0.001) | -0.007 (-0.016, 0.001) |
|  | ARI with Fever | Any antibiotic use in past month | -0.004 (-0.011, 0.000) | -0.008 (-0.017, -0.001) |
|  | Difficulty Breathing | Any antibiotic use in past month | 0.000 (-0.005, 0.003) | -0.004 (-0.011, 0.002) |
|  | Fever (Past 7 Days) | Any antibiotic use in past month | -0.003 (-0.013, 0.004) | -0.003 (-0.009, 0.002) |
|  | Fever (Past 14 Days) | Any antibiotic use in past month | -0.007 (-0.030, 0.013) | -0.009 (-0.035, 0.015) |
|  | Any Enteric Virus | Any antibiotic use in past month | -0.017 (-0.049, 0.008) | -0.018 (-0.040, -0.004) |
|  |  | Any antibiotic use in past 3 months | -0.007 (-0.031, 0.013) | -0.014 (-0.031, 0.002) |
|  | Any Enteric Virus with Diarrhea | Any antibiotic use in past month | -0.009 (-0.021, -0.001) | -0.013 (-0.025, -0.005) |
|  |  | Any antibiotic use in past 3 months | -0.008 (-0.021, 0.000) | -0.007 (-0.017, 0.000) |
|  |  | Episodes of antibiotic use in past 3 months | -0.023 (-0.053, 0.000) | -0.009 (-0.027, 0.005) |
|  |  | Multiple episodes of antibiotic use | -0.005 (-0.017, 0.003) | -0.004 (-0.015, 0.002) |
|  | Number of Enteric Viruses | Any antibiotic use in past month | -0.008 (-0.028, 0.009) | -0.008 (-0.021, 0.001) |
| Pooled Nutrition and Nutrition + WSH | Diarrhea, Fever, ARI, or Enteric Virus | Any antibiotic use in past month | -0.014 (-0.042, 0.002) | -0.013 (-0.035, 0.002) |
|  | Diarrhea, Fever, or ARI | Any antibiotic use in past month | -0.006 (-0.017, 0.001) | -0.005 (-0.013, 0.001) |
|  | Diarrhea | Any antibiotic use in past month | -0.006 (-0.015, 0.000) | -0.005 (-0.014, 0.000) |
|  | ARI | Any antibiotic use in past month | -0.002 (-0.008, 0.002) | -0.005 (-0.014, 0.002) |
|  | ARI with Fever | Any antibiotic use in past month | -0.003 (-0.009, 0.001) | -0.006 (-0.016, 0.002) |
|  | Difficulty Breathing | Any antibiotic use in past month | 0.000 (-0.005, 0.002) | -0.004 (-0.013, 0.003) |
|  | Fever (Past 7 Days) | Any antibiotic use in past month | -0.004 (-0.014, 0.003) | -0.002 (-0.008, 0.002) |
|  | Fever (Past 14 Days) | Any antibiotic use in past month | -0.010 (-0.033, 0.009) | -0.010 (-0.034, 0.009) |
|  | Any Enteric Virus | Any antibiotic use in past month | -0.015 (-0.048, 0.007) | -0.013 (-0.033, 0.001) |
|  |  | Any antibiotic use in past 3 months | -0.007 (-0.032, 0.009) | -0.007 (-0.024, 0.006) |
|  | Any Enteric Virus with Diarrhea | Any antibiotic use in past month | -0.006 (-0.017, 0.000) | -0.011 (-0.024, -0.003) |
|  |  | Any antibiotic use in past 3 months | -0.009 (-0.021, -0.001) | -0.007 (-0.018, 0.001) |
|  |  | Episodes of antibiotic use in past 3 months | -0.020 (-0.048, 0.000) | 0.000 (-0.017, 0.015) |
|  |  | Multiple episodes of antibiotic use | -0.005 (-0.017, 0.003) | 0.001 (-0.010, 0.007) |
|  | Number of Enteric Viruses | Any antibiotic use in past month | -0.008 (-0.027, 0.005) | -0.005 (-0.017, 0.005) |
| Pooled WSH and Nutrition + WSH | Diarrhea, Fever, ARI, or Enteric Virus | Any antibiotic use in past month | -0.021 (-0.057, 0.000) | -0.051 (-0.108, -0.011) |
|  | Diarrhea, Fever, or ARI | Any antibiotic use in past month | -0.008 (-0.020, 0.001) | -0.011 (-0.027, 0.002) |
|  | Diarrhea | Any antibiotic use in past month | -0.008 (-0.020, -0.001) | -0.009 (-0.019, -0.002) |
|  | ARI | Any antibiotic use in past month | -0.004 (-0.014, 0.001) | -0.011 (-0.025, 0.000) |
|  | ARI with Fever | Any antibiotic use in past month | -0.005 (-0.012, 0.000) | -0.010 (-0.022, -0.001) |
|  | Difficulty Breathing | Any antibiotic use in past month | -0.002 (-0.009, 0.003) | -0.007 (-0.017, 0.000) |
|  | Fever (Past 7 Days) | Any antibiotic use in past month | -0.005 (-0.015, 0.003) | -0.005 (-0.016, 0.003) |
|  | Fever (Past 14 Days) | Any antibiotic use in past month | -0.004 (-0.025, 0.016) | -0.005 (-0.036, 0.021) |
|  | Any Enteric Virus | Any antibiotic use in past month | -0.019 (-0.054, 0.008) | -0.026 (-0.057, -0.005) |
|  |  | Any antibiotic use in past 3 months | -0.006 (-0.030, 0.017) | -0.017 (-0.040, 0.001) |
|  | Any Enteric Virus with Diarrhea | Any antibiotic use in past month | -0.011 (-0.028, -0.001) | -0.016 (-0.035, -0.004) |
|  |  | Any antibiotic use in past 3 months | -0.008 (-0.020, 0.001) | -0.008 (-0.020, 0.001) |
|  |  | Episodes of antibiotic use in past 3 months | -0.022 (-0.052, 0.003) | -0.018 (-0.044, 0.002) |
|  |  | Multiple episodes of antibiotic use | -0.004 (-0.018, 0.005) | -0.010 (-0.028, 0.001) |
|  | Number of Enteric Viruses | Any antibiotic use in past month | -0.008 (-0.034, 0.013) | -0.017 (-0.034, -0.003) |
| Nutrition | Diarrhea, Fever, ARI, or Enteric Virus | Any antibiotic use in past month | -0.016 (-0.049, 0.006) | -0.010 (-0.034, 0.004) |
|  | Diarrhea, Fever, or ARI | Any antibiotic use in past month | -0.002 (-0.013, 0.007) | -0.001 (-0.009, 0.005) |
|  | Diarrhea | Any antibiotic use in past month | -0.003 (-0.013, 0.003) | -0.002 (-0.011, 0.003) |
|  | ARI | Any antibiotic use in past month | -0.001 (-0.007, 0.002) | -0.003 (-0.016, 0.007) |
|  | ARI with Fever | Any antibiotic use in past month | -0.003 (-0.010, 0.001) | -0.007 (-0.021, 0.002) |
|  | Difficulty Breathing | Any antibiotic use in past month | 0.000 (-0.004, 0.002) | -0.002 (-0.010, 0.006) |
|  | Fever (Past 7 Days) | Any antibiotic use in past month | -0.003 (-0.014, 0.005) | -0.002 (-0.008, 0.003) |
|  | Fever (Past 14 Days) | Any antibiotic use in past month | -0.008 (-0.032, 0.009) | -0.013 (-0.050, 0.016) |
|  | Any Enteric Virus | Any antibiotic use in past month | -0.016 (-0.049, 0.009) | -0.014 (-0.042, 0.005) |
|  |  | Any antibiotic use in past 3 months | -0.007 (-0.035, 0.014) | -0.005 (-0.032, 0.017) |
|  | Any Enteric Virus with Diarrhea | Any antibiotic use in past month | -0.005 (-0.017, 0.001) | -0.013 (-0.029, -0.003) |
|  |  | Any antibiotic use in past 3 months | -0.008 (-0.020, -0.001) | -0.006 (-0.021, 0.004) |
|  |  | Episodes of antibiotic use in past 3 months | -0.013 (-0.035, 0.000) | 0.002 (-0.011, 0.018) |
|  |  | Multiple episodes of antibiotic use | -0.005 (-0.018, 0.002) | 0.001 (-0.011, 0.009) |
|  | Number of Enteric Viruses | Any antibiotic use in past month | -0.009 (-0.028, 0.006) | -0.002 (-0.019, 0.013) |
| WSH | Diarrhea, Fever, ARI, or Enteric Virus | Any antibiotic use in past month | -0.022 (-0.062, 0.002) | -0.098 (-0.246, -0.005) |
|  | Diarrhea, Fever, or ARI | Any antibiotic use in past month | -0.003 (-0.015, 0.006) | -0.007 (-0.029, 0.014) |
|  | Diarrhea | Any antibiotic use in past month | -0.006 (-0.018, 0.000) | -0.009 (-0.024, -0.001) |
|  | ARI | Any antibiotic use in past month | -0.002 (-0.010, 0.003) | -0.009 (-0.028, 0.006) |
|  | ARI with Fever | Any antibiotic use in past month | -0.004 (-0.012, 0.001) | -0.011 (-0.028, 0.001) |
|  | Difficulty Breathing | Any antibiotic use in past month | -0.001 (-0.008, 0.003) | -0.003 (-0.012, 0.002) |
|  | Fever (Past 7 Days) | Any antibiotic use in past month | -0.003 (-0.014, 0.006) | -0.004 (-0.020, 0.009) |
|  | Fever (Past 14 Days) | Any antibiotic use in past month | 0.001 (-0.023, 0.026) | 0.003 (-0.042, 0.046) |
|  | Any Enteric Virus | Any antibiotic use in past month | -0.018 (-0.060, 0.017) | -0.038 (-0.087, -0.004) |
|  |  | Any antibiotic use in past 3 months | -0.009 (-0.041, 0.019) | -0.029 (-0.070, 0.001) |
|  | Any Enteric Virus with Diarrhea | Any antibiotic use in past month | -0.010 (-0.025, 0.000) | -0.017 (-0.044, -0.003) |
|  |  | Any antibiotic use in past 3 months | -0.008 (-0.020, 0.000) | -0.008 (-0.024, 0.002) |
|  |  | Episodes of antibiotic use in past 3 months | -0.022 (-0.056, 0.002) | -0.029 (-0.069, -0.002) |
|  |  | Multiple episodes of antibiotic use | -0.005 (-0.018, 0.004) | -0.019 (-0.054, -0.003) |
|  | Number of Enteric Viruses | Any antibiotic use in past month | -0.008 (-0.039, 0.019) | -0.023 (-0.055, 0.001) |
| Nutrition + WSH | Diarrhea, Fever, ARI, or Enteric Virus | Any antibiotic use in past month | -0.016 (-0.048, 0.006) | -0.020 (-0.058, 0.008) |
|  | Diarrhea, Fever, or ARI | Any antibiotic use in past month | -0.011 (-0.027, -0.001) | -0.010 (-0.026, 0.000) |
|  | Diarrhea | Any antibiotic use in past month | -0.008 (-0.021, -0.001) | -0.010 (-0.028, -0.001) |
|  | ARI | Any antibiotic use in past month | -0.003 (-0.011, 0.002) | -0.007 (-0.019, 0.001) |
|  | ARI with Fever | Any antibiotic use in past month | -0.003 (-0.011, 0.001) | -0.005 (-0.017, 0.003) |
|  | Difficulty Breathing | Any antibiotic use in past month | -0.001 (-0.006, 0.003) | -0.007 (-0.020, 0.003) |
|  | Fever (Past 7 Days) | Any antibiotic use in past month | -0.005 (-0.016, 0.004) | -0.003 (-0.013, 0.003) |
|  | Fever (Past 14 Days) | Any antibiotic use in past month | -0.008 (-0.035, 0.016) | -0.008 (-0.035, 0.015) |
|  | Any Enteric Virus | Any antibiotic use in past month | -0.013 (-0.048, 0.007) | -0.013 (-0.039, 0.005) |
|  |  | Any antibiotic use in past 3 months | -0.007 (-0.032, 0.011) | -0.013 (-0.040, 0.006) |
|  | Any Enteric Virus with Diarrhea | Any antibiotic use in past month | -0.007 (-0.020, 0.000) | -0.012 (-0.032, 0.000) |
|  |  | Any antibiotic use in past 3 months | -0.008 (-0.019, 0.000) | -0.009 (-0.031, 0.005) |
|  |  | Episodes of antibiotic use in past 3 months | -0.022 (-0.053, 0.003) | -0.004 (-0.036, 0.022) |
|  |  | Multiple episodes of antibiotic use | -0.003 (-0.016, 0.006) | -0.001 (-0.025, 0.011) |
|  | Number of Enteric Viruses | Any antibiotic use in past month | -0.007 (-0.024, 0.007) | -0.008 (-0.026, 0.004) |

Point estimates and 95% confidence intervals for the total and pure natural indirect effects of WASH and/or Nutrition intervention on antibiotic use, computed only when the difference in total and pure natural indirect effects was greater than 1% or had a t-test p-value < 0.2 when outcome models included an intervention-mediator interaction term. For categorical outcomes (any antibiotic use, multiple episodes of antibiotic use), the prevalence difference is reported. For continuous outcomes (days of antibiotic use, episodes of antibiotic use), the mean difference is reported. Diarrhea, ARI, ARI with Fever, and Fever are all reported by a caregiver under a 7-day lookback period at 14 and 28 months. Any Enteric Virus is the presence of adenovirus 40/41, norovirus GI, norovirus GII, sapovirus, rotavirus, or astrovirus in stool collected at 14 months, and Any Enteric Virus with Diarrhea is the presence of any enteric virus with caregiver reported diarrhea in the prior 7 days at 14 months. Antibiotic use is reported by a caregiver under either a 1- or 3-month lookback period at 14 and 28 months.

### Table S10: Intervention-Mediator Effects, for Enteric Viruses with Pathogen Loads Reflecting Diarrheal Etiology

| **Intervention Group** | **Mediator** | **Prevalence/Mean in Control Arm** | **Prevalence/Mean in Intervention Arm** | **Prevalence Ratio (95% CI)** |
| --- | --- | --- | --- | --- |
| Pooled Intervention | Any Enteric Virus | 0.401 (0.332, 0.469) | 0.217 (0.184, 0.250) | 0.516 ( 0.411, 0.648) |
|  | Adenovirus 40/41 | 0.101 (0.049, 0.152) | 0.068 (0.046, 0.090) | 0.593 ( 0.357, 0.983) |
|  | Norovirus GII | 0.164 (0.119, 0.209) | 0.098 (0.077, 0.120) | 0.578 ( 0.397, 0.841) |
|  | Sapovirus | 0.166 (0.119, 0.214) | 0.078 (0.060, 0.096) | 0.454 ( 0.305, 0.677) |
| Pooled Nutrition and Nutrition + WSH | Any Enteric Virus | 0.401 (0.332, 0.469) | 0.247 (0.205, 0.290) | 0.595 ( 0.467, 0.758) |
|  | Adenovirus 40/41 | 0.101 (0.049, 0.152) | 0.074 (0.045, 0.103) | 0.660 ( 0.393, 1.107) |
|  | Norovirus GII | 0.164 (0.119, 0.209) | 0.120 (0.090, 0.149) | 0.718 ( 0.487, 1.058) |
|  | Sapovirus | 0.166 (0.119, 0.214) | 0.084 (0.062, 0.106) | 0.494 ( 0.319, 0.763) |
| Pooled WSH and Nutrition + WSH | Any Enteric Virus | 0.401 (0.332, 0.469) | 0.204 (0.171, 0.237) | 0.486 ( 0.381, 0.619) |
|  | Adenovirus 40/41 | 0.101 (0.049, 0.152) | 0.061 (0.040, 0.083) | 0.506 ( 0.282, 0.909) |
|  | Norovirus GII | 0.164 (0.119, 0.209) | 0.088 (0.066, 0.110) | 0.519 ( 0.343, 0.786) |
|  | Sapovirus | 0.166 (0.119, 0.214) | 0.078 (0.057, 0.099) | 0.463 ( 0.306, 0.701) |
| Nutrition | Any Enteric Virus | 0.401 (0.332, 0.469) | 0.244 (0.181, 0.308) | 0.569 ( 0.421, 0.769) |
|  | Adenovirus 40/41 | 0.101 (0.049, 0.152) | 0.080 (0.042, 0.119) | 0.724 ( 0.403, 1.300) |
|  | Norovirus GII | 0.164 (0.119, 0.209) | 0.119 (0.074, 0.164) | 0.705 ( 0.441, 1.129) |
|  | Sapovirus | 0.166 (0.119, 0.214) | 0.077 (0.044, 0.110) | 0.421 ( 0.235, 0.753) |
| WSH | Any Enteric Virus | 0.401 (0.332, 0.469) | 0.159 (0.114, 0.204) | 0.359 ( 0.258, 0.499) |
|  | Adenovirus 40/41 | 0.101 (0.049, 0.152) | 0.055 (0.030, 0.081) | 0.439 ( 0.218, 0.883) |
|  | Norovirus GII | 0.164 (0.119, 0.209) | 0.057 (0.033, 0.081) | 0.321 ( 0.190, 0.544) |
|  | Sapovirus | 0.166 (0.119, 0.214) | 0.066 (0.035, 0.098) | 0.363 ( 0.202, 0.653) |
| Nutrition + WSH | Any Enteric Virus | 0.401 (0.332, 0.469) | 0.250 (0.197, 0.303) | 0.607 ( 0.457, 0.806) |
|  | Adenovirus 40/41 | 0.101 (0.049, 0.152) | 0.068 (0.036, 0.100) | 0.567 ( 0.301, 1.067) |
|  | Norovirus GII | 0.164 (0.119, 0.209) | 0.120 (0.081, 0.159) | 0.727 ( 0.456, 1.159) |
|  | Sapovirus | 0.166 (0.119, 0.214) | 0.090 (0.060, 0.121) | 0.546 ( 0.339, 0.878) |

Prevalance ratio estimates and 95% confidence intervals for the effects of WASH and/or Nutrition interventions on enteric virus carriage at 14 months with pathogen loads that reflect diarrheal etiology based on published Ct cutoff values from the MAL-ED study. Any Enteric Virus is the presence of Adenovirus 40/41, Norovirus GII, or Sapovirus that exceeded the etiology cutoff.

### Table S11: Mediator-Outcome Effects, for Enteric Viruses with Pathogen Loads Reflecting Diarrheal Etiology

| **Mediator** | **Outcome** | **Prevalence/Mean in Absence of Mediator** | **Prevalence/Mean in Presence of Mediator** | **Mean Difference (95% CI)** | **Prevalence Ratio (95% CI)** |
| --- | --- | --- | --- | --- | --- |
| Any Enteric Virus | Any antibiotic use in past month | 0.535 (0.502, 0.567) | 0.618 (0.568, 0.667) | ---- | 1.141 ( 1.025, 1.270) |
|  | Any antibiotic use in past 3 months | 0.711 (0.680, 0.743) | 0.737 (0.686, 0.788) | ---- | 1.042 ( 0.955, 1.137) |
|  | Days of antibiotic use in past 3 months | 5.607 (5.279, 5.936) | 5.750 (5.104, 6.396) | 0.130 (-0.600, 0.861) | ---- |
|  | Episodes of antibiotic use in past 3 months | 1.088 (1.027, 1.149) | 1.178 (1.066, 1.289) | 0.076 (-0.061, 0.213) | ---- |
|  | Multiple episodes of antibiotic use | 0.270 (0.243, 0.297) | 0.325 (0.271, 0.379) | ---- | 1.167 ( 0.963, 1.415) |
| Adenovirus 40/41 | Any antibiotic use in past month | 0.558 (0.529, 0.587) | 0.552 (0.455, 0.650) | ---- | 0.993 ( 0.826, 1.193) |
|  | Any antibiotic use in past 3 months | 0.720 (0.694, 0.747) | 0.714 (0.646, 0.783) | ---- | 0.990 ( 0.887, 1.106) |
|  | Days of antibiotic use in past 3 months | 5.694 (5.385, 6.002) | 5.317 (4.418, 6.217) | -0.292 (-1.256, 0.671) | ---- |
|  | Episodes of antibiotic use in past 3 months | 1.107 (1.055, 1.159) | 1.181 (0.989, 1.373) | 0.075 (-0.125, 0.275) | ---- |
|  | Multiple episodes of antibiotic use | 0.282 (0.257, 0.307) | 0.314 (0.210, 0.419) | ---- | 1.126 ( 0.817, 1.552) |
| Norovirus GII | Any antibiotic use in past month | 0.550 (0.523, 0.577) | 0.631 (0.560, 0.702) | ---- | 1.125 ( 0.992, 1.276) |
|  | Any antibiotic use in past 3 months | 0.721 (0.696, 0.747) | 0.725 (0.664, 0.786) | ---- | 1.000 ( 0.910, 1.099) |
|  | Days of antibiotic use in past 3 months | 5.667 (5.371, 5.962) | 5.742 (4.829, 6.656) | -0.033 (-0.979, 0.914) | ---- |
|  | Episodes of antibiotic use in past 3 months | 1.113 (1.061, 1.164) | 1.131 (0.995, 1.267) | -0.016 (-0.166, 0.133) | ---- |
|  | Multiple episodes of antibiotic use | 0.281 (0.256, 0.306) | 0.306 (0.235, 0.378) | ---- | 1.035 ( 0.813, 1.317) |
| Sapovirus | Any antibiotic use in past month | 0.551 (0.523, 0.578) | 0.627 (0.546, 0.708) | ---- | 1.129 ( 0.983, 1.297) |
|  | Any antibiotic use in past 3 months | 0.717 (0.691, 0.742) | 0.748 (0.667, 0.830) | ---- | 1.054 ( 0.942, 1.179) |
|  | Days of antibiotic use in past 3 months | 5.624 (5.312, 5.937) | 5.933 (4.792, 7.074) | 0.354 (-0.806, 1.514) | ---- |
|  | Episodes of antibiotic use in past 3 months | 1.106 (1.050, 1.162) | 1.185 (1.014, 1.356) | 0.076 (-0.108, 0.260) | ---- |
|  | Multiple episodes of antibiotic use | 0.278 (0.252, 0.305) | 0.333 (0.246, 0.421) | ---- | 1.175 ( 0.884, 1.561) |

Point estimates and 95% confidence intervals for the effect of enteric virus carriage at 14 months with pathogen loads that reflect diarrheal etiology. For categorical mediators, the prevalence ratio is reported. For continuous mediators, the mean difference is reported. Diarrheal etiology is assessed using published Ct cutoff values from the MAL-ED study. Any Enteric Virus is the presence of Adenovirus 40/41, Norovirus GII, or Sapovirus that exceeded the etiology cutoff. Antibiotic use is reported by a caregiver under either a 1- or 3-month lookback period at 14 and 28 months.

### Table S12: Mediated Effects, for Enteric Viruses with Pathogen Loads Reflecting Diarrheal Etiology

| **Intervention Group** | **Mediator** | **Outcome** | **Total Natural Indirect Effect (Prevalence Difference)** | **Total Natural Indirect Effect (Prevalence Ratio)** |
| --- | --- | --- | --- | --- |
| Pooled Intervention | Any Enteric Virus | Any antibiotic use in past month | -0.012 (-0.030, -0.002) | 0.985 (0.968, 0.998) |
| Pooled Nutrition and Nutrition + WSH | Any Enteric Virus | Any antibiotic use in past month | -0.006 (-0.020, 0.002) | 0.993 (0.977, 1.006) |
| Pooled WSH and Nutrition + WSH | Any Enteric Virus | Any antibiotic use in past month | -0.022 (-0.046, -0.005) | 0.975 (0.952, 0.992) |
| Nutrition | Any Enteric Virus | Any antibiotic use in past month | -0.004 (-0.021, 0.008) | 0.995 (0.974, 1.018) |
| WSH | Any Enteric Virus | Any antibiotic use in past month | -0.028 (-0.061, -0.002) | 0.965 (0.933, 0.996) |
| Nutrition + WSH | Any Enteric Virus | Any antibiotic use in past month | -0.010 (-0.027, 0.001) | 0.991 (0.973, 1.006) |

Point estimates and 95% confidence intervals for the total natural indirect effects of WASH and/or Nutrition intervention on antibiotic use through pathways involving enteric virus carriage at 14 months with pathogen loads that reflect diarrheal etiology. Diarrheal etiology is assessed using published Ct cutoff values from the MAL-ED study. Any Enteric Virus is the presence of Adenovirus 40/41, Norovirus GII, or Sapovirus that exceeded the etiology cutoff. Antibiotic use is reported by a caregiver under either a 1- or 3-month lookback period at 14 and 28 months.

### Table S13: Mediated Effects, Negative Control

| **Intervention Group** | **Mediator** | **Outcome** | **Total Natural Indirect Effect (Prevalence Difference)** |
| --- | --- | --- | --- |
| Pooled Intervention | Bruising (Past 7 Days) | Any antibiotic use in past month | -0.001 (-0.004, 0.001) |
|  |  | Any antibiotic use in past 3 months | -0.002 (-0.006, 0.001) |
|  |  | Days of antibiotic use in past 3 months | -0.006 (-0.035, 0.018) |
|  |  | Episodes of antibiotic use in past 3 months | -0.001 (-0.006, 0.004) |
|  |  | Multiple episodes of antibiotic use | 0.000 (-0.001, 0.002) |
| Pooled Nutrition and Nutrition + WSH | Bruising (Past 7 Days) | Any antibiotic use in past month | 0.000 (-0.003, 0.002) |
|  |  | Any antibiotic use in past 3 months | -0.001 (-0.005, 0.002) |
|  |  | Days of antibiotic use in past 3 months | -0.006 (-0.047, 0.032) |
|  |  | Episodes of antibiotic use in past 3 months | 0.000 (-0.005, 0.004) |
|  |  | Multiple episodes of antibiotic use | 0.000 (-0.002, 0.002) |
| Pooled WSH and  Nutrition + WSH | Bruising (Past 7 Days) | Any antibiotic use in past month | -0.001 (-0.007, 0.002) |
|  |  | Any antibiotic use in past 3 months | -0.002 (-0.008, 0.001) |
|  |  | Days of antibiotic use in past 3 months | -0.016 (-0.066, 0.014) |
|  |  | Episodes of antibiotic use in past 3 months | 0.000 (-0.006, 0.005) |
|  |  | Multiple episodes of antibiotic use | 0.001 (-0.001, 0.004) |
| Nutrition | Bruising (Past 7 Days) | Any antibiotic use in past month | 0.000 (-0.003, 0.004) |
|  |  | Any antibiotic use in past 3 months | -0.001 (-0.007, 0.004) |
|  |  | Days of antibiotic use in past 3 months | -0.012 (-0.090, 0.052) |
|  |  | Episodes of antibiotic use in past 3 months | 0.000 (-0.011, 0.009) |
|  |  | Multiple episodes of antibiotic use | 0.000 (-0.004, 0.003) |
| WSH | Bruising (Past 7 Days) | Any antibiotic use in past month | -0.004 (-0.011, 0.001) |
|  |  | Any antibiotic use in past 3 months | -0.003 (-0.010, 0.001) |
|  |  | Days of antibiotic use in past 3 months | -0.029 (-0.099, 0.011) |
|  |  | Episodes of antibiotic use in past 3 months | -0.002 (-0.011, 0.006) |
|  |  | Multiple episodes of antibiotic use | 0.002 (-0.001, 0.005) |
| Nutrition + WSH | Bruising (Past 7 Days) | Any antibiotic use in past month | 0.000 (-0.003, 0.002) |
|  |  | Any antibiotic use in past 3 months | -0.001 (-0.006, 0.002) |
|  |  | Days of antibiotic use in past 3 months | -0.013 (-0.075, 0.028) |
|  |  | Episodes of antibiotic use in past 3 months | 0.000 (-0.006, 0.007) |
|  |  | Multiple episodes of antibiotic use | 0.000 (-0.003, 0.003) |

Point estimates and 95% confidence intervals for the total natural indirect effects of WASH and/or Nutrition intervention on antibiotic use through pathways involving bruising, a negative control. Bruising is reported by a caregiver under a 7-day lookback period at 14 and 28 months. Antibiotic use is reported by a caregiver under either a 1- or 3-month lookback period at 14 and 28 months.
